# COVID-19 pandemic in Saint Petersburg, Russia: combining surveillance and population-based serological study data in May, 2020–April, 2021

**DOI:** 10.1101/2021.07.31.21261428

**Authors:** Anton Barchuk, Dmitriy Skougarevskiy, Alexei Kouprianov, Daniil Shirokov, Olga Dudkina, Rustam Tursun-zade, Mariia Sergeeva, Varvara Tychkova, Andrey Komissarov, Alena Zheltukhina, Dmitry Lioznov, Artur Isaev, Ekaterina Pomerantseva, Svetlana Zhikrivetskaya, Yana Sofronova, Konstantin Blagodatskikh, Kirill Titaev, Lubov Barabanova, Daria Danilenko

## Abstract

**Background:** The COVID-19 pandemic in Russia has already resulted in 500,000 excess deaths, with more than 5.6 million cases registered officially by July 2021. Surveillance based on case reporting has become the core pandemic monitoring method in the country and globally. However, population-based seroprevalence studies may provide an unbiased estimate of the actual disease spread and, in combination with multiple surveillance tools, help to define the pandemic course. This study summarises results from four consecutive serological surveys conducted between May 2020 and April 2021 at St. Petersburg, Russia and combines them with other SARS-CoV-2 surveillance data.

**Methods:** We conducted four serological surveys of two random samples (May–June, July–August, October–December 2020, and February–April 2021) from adults residing in St. Petersburg recruited with the random digit dialing (RDD), accompanied by a telephone interview to collect information on both individuals who accepted and declined the invitation for testing and account for non-response. We have used enzyme-linked immunosorbent assay CoronaPass total antibodies test (Genetico, Moscow, Russia) to report seroprevalence. We corrected the estimates for non-response using the bivariate probit model and also accounted the test performance characteristics, obtained from independent assay evaluation. In addition, we have summarised the official registered cases statistics, the number of hospitalised patients, the number of COVID-19 deaths, excess deaths, tests performed, data from the ongoing SARS-CoV-2 variants of concern (VOC) surveillance, the vaccination uptake, and St. Petersburg search and mobility trends. The infection fatality ratios (IFR) have been calculated using the Bayesian evidence synthesis model.

**Findings:** After calling 113,017 random mobile phones we have reached 14,118 individuals who responded to computer-assisted telephone interviewing (CATI) and 2,413 provided blood samples at least once through the seroprevalence study. The adjusted seroprevalence in May–June, 2020 was 9.7% (95%: 7.7–11.7), 13.3% (95% 9.9–16.6) in July–August, 2020, 22.9% (95%: 20.3–25.5) in October–December, 2021 and 43.9% (95%: 39.7–48.0) in February–April, 2021. History of any symptoms, history of COVID-19 tests, and non-smoking status were significant predictors for higher seroprevalence. Most individuals remained seropositive with a maximum 10 months follow-up. 92.7% (95% CI 87.9–95.7) of participants who have reported at least one vaccine dose were seropositive. Hospitalisation and COVID-19 death statistics and search terms trends reflected the pandemic course better than the official case count, especially during the spring 2020. SARS-CoV-2 circulation showed rather low genetic SARS-CoV-2 lineages diversity that increased in the spring 2021. Local VOC (AT.1) was spreading till April 2021, but B.1.617.2 substituted all other lineages by June 2021. The IFR based on the excess deaths was equal to 1.04 (95% CI 0.80–1.31) for the adult population and 0.86% (95% CI 0.66–1.08) for the entire population.

**Conclusion:** Approximately one year after the COVID-19 pandemic about 45% of St. Petersburg, Russia residents contracted the SARS-CoV-2 infection. Combined with vaccination uptake of about 10% it was enough to slow the pandemic until the Delta VOC started to spread. Combination of several surveillance tools provides a comprehensive pandemic picture.

**Funding:** Polymetal International plc.

## Introduction

The COVID-19 pandemic in Russia has already resulted in 500,000 excess deaths [1], with more than 5.6 million cases registered officially by July 2021 [2]. Surveillance based on case reporting has become the core method for monitoring the pandemic in Russia and globally. However, the actual spread of SARS-CoV-2 is challenging to measure as case definitions, testing strategies, and capacity are not comparable between countries and in the different periods [3]. Population-based studies using representative samples of the population combined with serological assessment for the presence of SARS-CoV-2 may provide an unbiased estimate of the actual disease spread and help estimate the true disease burden as well as the infection fatality rate (IFR) [4–7]. Unfortunately, national serological studies to assess the prevalence of SARS-CoV-2 in Russia were not yet published. Given a considerable territory, it is not likely that the pandemic develops similarly across the country. Therefore, different studies are needed to explore seroprevalence and the pandemic course in big cities and less densely populated regions. Saint Petersburg is the second-largest city in Russia, with the first SARS-CoV-2 case registered on March 5, 2020. Seroprevalence study conducted in St. Petersburg between May 27 and June 26, 2020, estimated that not more than 10% of the population had contracted the SARS-CoV-2 [8]. These findings were in line with seroprevalence estimates in other European studies summarised in the systematic review [9], which revealed only 82 studies of higher quality out of 404 studies included in the meta-analyses. Lack of sample representativeness and methods to correct participants’ characteristics and test performance limited the quality for most assessed studies.

Official case count and serological studies are not the only methods for SARS-CoV-2 surveillance. Cause-specific COVID-19 mortality is another valuable statistic to assess the pandemic impact. However, it may be biased in different healthcare settings, especially when definitions for COVID-19 death are not comparable. Using excess mortality, i.e. quantifying deaths from all causes relative to a recent historical benchmark, can help avoid this bias [1, 10]. St. Petersburg was one of the two Russian regions with the most reliable reporting of COVID-19 mortality [11]. SARS-CoV-2 variants of concern (VOC) monitoring is another critical surveillance tool that turned to become crucial in the later stages of the COVID-19 pandemic when new, more transmissive VOCs started to spread rapidly [12, 13].

Novel auxiliary surveillance methods like search term trends to monitor the COVID-19 pandemic and mobility trends to monitor the effects of mitigation measures and population behaviour can also be helpful [14, 15]. For example, search terms and mobility trends are available for St. Petersburg. However, these low-barrier research methods are often criticised for the lack of validity [16]. This study summarises the four consecutive rounds of population-based serological study based on two representative samples of adults residing in St. Petersburg, Russia, between May 2020 and April 2021. In addition, we combine the seroprevalence estimates with all other available surveillance data: official case count, hospitalisation data, SARS-CoV-2 VOCs monitoring data, COVID-19 specific mortality, excess mortality, vaccination uptake, mobility trends, and search term trends. Thus, we aim to assess the different surveillance tools validity and present a comprehensive pandemic course in the fourth largest European city with a more than 5 million population.

## Materials and methods

### Seroprevalence of anti-SARS-CoV-2 antibodies

St. Petersburg serological study settings and design are described in detail in our previous report [8]. In brief, St. Petersburg COVID-19 study is population-based epidemiological survey of a random sample from the adult population to assess the sero-prevalence of anti-SARS-CoV-2 antibodies. The study was based on a phone-based survey and an individual invitation to the clinic for blood sample collection. Eligible individuals were adults residing in St. Petersburg older than 18 years and recruited using the random digit dialling (RDD) method. RDD was accompanied by the computer-assisted telephone interviewing (CATI) to collect the information on both individuals who accepted and declined the invitation for testing.

Blood samples from the same population group were collected between May 25, 2020, and June 28, 2020, in the first cross-section “May–June 2020 survey” henceforth) and between July 20, 2020, and August 8, 2020, in the second “July–August 2020 survey” cross-section). Considering the risks of low response in the next planned cross-section, we created a new population sample applying the similar strategy of RDD followed by CATI (“October–December 2020 survey”). The initial response to the RDD was higher in autumn and winter 2020–2021 compared to the first cross-section in summer 2020. The fourth cross-section (“February–April 2021 survey”) involved individuals from both population samples invited between February 15, 2021, and April 4, 2021. Repeated blood sampling allowed seroconversion assessment for individuals who tested positive in previous surveys. Also, in this cross-section, some participants reported at least one vaccine shot. They were included in the study as non-responders as the initial survey does not fully address the characteristics associated with vaccination status. However, the vaccinated individuals were still tested. The participant flow for all four cross-sections is reported in Figure 1. The full study protocol is available online (https://eusp.org/sites/defult/files/inline-files/EU_SG-Russian-covid-serosurvey-Protocal-CDRU-001_en.pdf).

**Figure 1.**
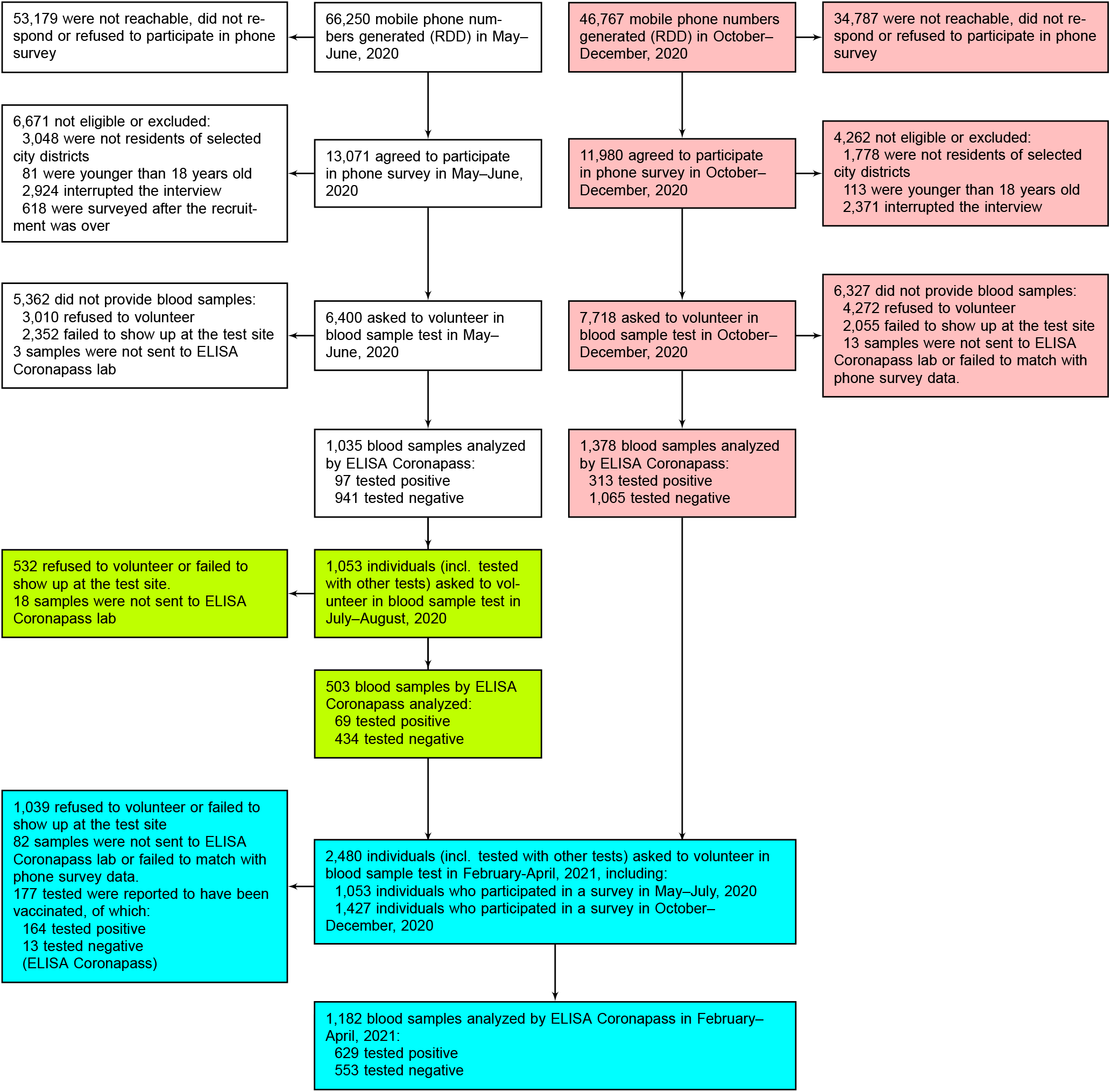
Flow chart of participants’ progress through the St. Petersburg seroprevalence study. Color codes study cross-sections: white — cross-section 1 (2020-05-25 – 2020-06-28), green — cross-section 2 (2020-07-20 – 2020-08-08), pink — cross-section 3 (2020-10-12 – 2020-12-06), blue — cross-section 4 (2020-02-15 – 2020-04-04).

### Laboratory tests

During the four surveys, we assessed anti-SARS-CoV-2 antibodies using three different assays. Even though our report was selected among studies of higher quality in the recent systematic review, a significant limitation was related to the absence of own test performance validation [9]. We conducted a validation that revealed the decrease in sensitivity for one of the assays [17]. Finally, to report seroprevalence, we selected enzyme-linked immunosorbent assay (ELISA Coronapass) CoronaPass total antibodies test (Genetico, Moscow, Russia) that detects total antibodies (the cutoff for positivity 1.0) and is based on the recombinant SARS-CoV-2 spike protein receptor binding domain (Department of Microbiology, Icahn School of Medicine at Mount Sinai, New York, NY, USA). We used ELISA Coronapass through all four surveys. We also used the results of our validation study to correct the seroprevalence estimate for test performance. Sensitivity is equal to 92% and specificity to 100% for ELISA Coronapass (for full validation see [17]).

### Surveillance data related to SARS-CoV-2 monitoring

We summarised the data that included the official registered cases statistics, the number of patients hospitalised, the number of COVID-19 deaths, excess deaths, and tests performed for COVID-19 detection. Although this information was not available from one source, we used a combination of different sources to restore the pandemic course in St. Petersburg. We have also used the leading Russian search engine Yandex search history in St. Petersburg region to obtain search trends for three terms: “loss of smell”, “smell”, and “saturation”. In addition, Yandex provided mobility trends for St. Petersburg from the open data from Yandex, Apple, and Otomono (https://yandex.ru/company/reserches/2020/cities-activity). Finally, we obtained data from the ongoing the Smorodintsev Research Institute of Influenza SARS-CoV-2 VOCs surveillance in St. Petersburg [18]. Data sources are described in detail in our Supplementary material.

### Infection fatality ratios

We used the information on the official COVID-19 mortality and derived excess mortality to estimate the IFR. IFR was calculated for the four periods covered by our seroprevalence surveys. We treated the true number of deaths as an interval censored random variable bound downwards/upwards by the number of deaths 14 days after the cross-section start/end date (see Supplementary Materials).

### Statistical analysis

The sample size calculations and statistical analysis plan for the serological survey were described in detail in our previous report [8]. The initial sample size of 1550 participants was calculated assuming the hypothetical prevalence of 20% to obtain the resulting sampling error of 2% using a 95% confidence interval. The actual sample size was lower, which resulted in a maximum error of about 4% when hypothetical seroprevalence reached 40%. The study’s primary aim was to assess the seroprevalence based on antibody tests accounting for non-response bias and test sensitivity and specificity. Non-response was evaluated by comparing answers provided during the CATI by those who visited the test site and all surveyed individuals, estimated using a binomial probit regression of individual agreement to participate in the study and offer their blood sample on their observable characteristics. In the first report, we described the variables that we had chosen to estimate the correction. The observable characteristics associated with response and positivity were reported any disease symptoms before the test and the COVID-19 testing history. We used similar variables to correct the seroprevalence estimates for non-response during all four cross-sections. To account possible sample non-representativeness in a sensitivity analysis, we computed raking weights to match the survey age group and educational attainment proportions in the 2016 representative survey of the adult city population with R package `anesrake` used to compute the weights. The original report also explored individual risk factors for test positivity in the sample participants who completed clinic paper-based surveys. This report assessed individual risk factors using a binomial probit regression used to estimate seroprevalence. Standard errors were computed with the delta method. For IFR computations we relied on a Bayesian evidence synthesis model [19] described in Supplementary Materials.

### Ethical considerations

The Research Planning Board approved the study of the European University at St. Petersburg (on May 20, 2020) and the Ethics Committee of the Clinic “Scandinavia” (on May 26, 2020). All research was performed following the relevant guidelines and regulations. Informed consent was obtained from all participants of the study. The study was registered with the following identifiers: Clinicaltrials.gov (NCT04406038, submitted on May 26, 2020, date of registration — May 28, 2020) and ISRCTN registry (ISRCTN11060415, submitted on May 26, 2020, date of registration — May 28, 2020). Official statistics, VOCs monitoring data, search terms trends, and mobility trends were obtained from open sources as aggregated data. Analysis based on open-source aggregated data does not require additional ethical permission in Russia.

### Data sharing

All analyses were conducted in _ with the aid of GJRM package [20], study data and code is available online (https://github.com/eusporg/spb_covid_study20)

### Role of the funding source

The study’s funder had no role in study design, data collection, data analysis, data interpretation, or writing of the report.

## Results

### Seroprevalence of antibodies to SARS-CoV-2

The resulting 14,118 individuals responded to CATI questionnaire — 6,400 in the first population sampling and 7,718 in the second (see Supplementary Appendix Table S5 for details regarding missing records on variables of interest). The respondents represent city population in terms of their gender, employment status, and household size, but were younger than the adult city population as of 2016 and had higher levels of educational attainment (see Supplementary Appendix Table S6). Overall, 2,413 individuals provided blood samples through the seroprevalence study course that were analysed using ELISA Coronapass: 1,035 in the first May–June 2020 survey and 503 of them in the second July–August survey, and 1,378 newly recruited participants in the third October–December survey. Finally, samples from 1,182 participants from previous surveys were collected and analysed in February–April 2021.

The adjusted seroprevalence in May–June 2020 was 9.7% (95%: 7.7–11.7) and increased to 13.3% (95% 9.9–16.6) in July– August 2020. We noticed a major increase through the third (22.9% 95%: 20.3–25.5) and between the third and fourth cross-sections of the seroprevalence study (see Figure 3 and Supplementary Figure S1 for the weekly data), resulting in seroprevalence equal to 43.9% (95%: 39.7–48.0) in February–April 2021. Naïve antibodies seroprevalence to SARS-CoV-2 and seroprevalence corrected for non-response only and corrected for non-response and test performance are presented in Table 1.

**Table 1.**
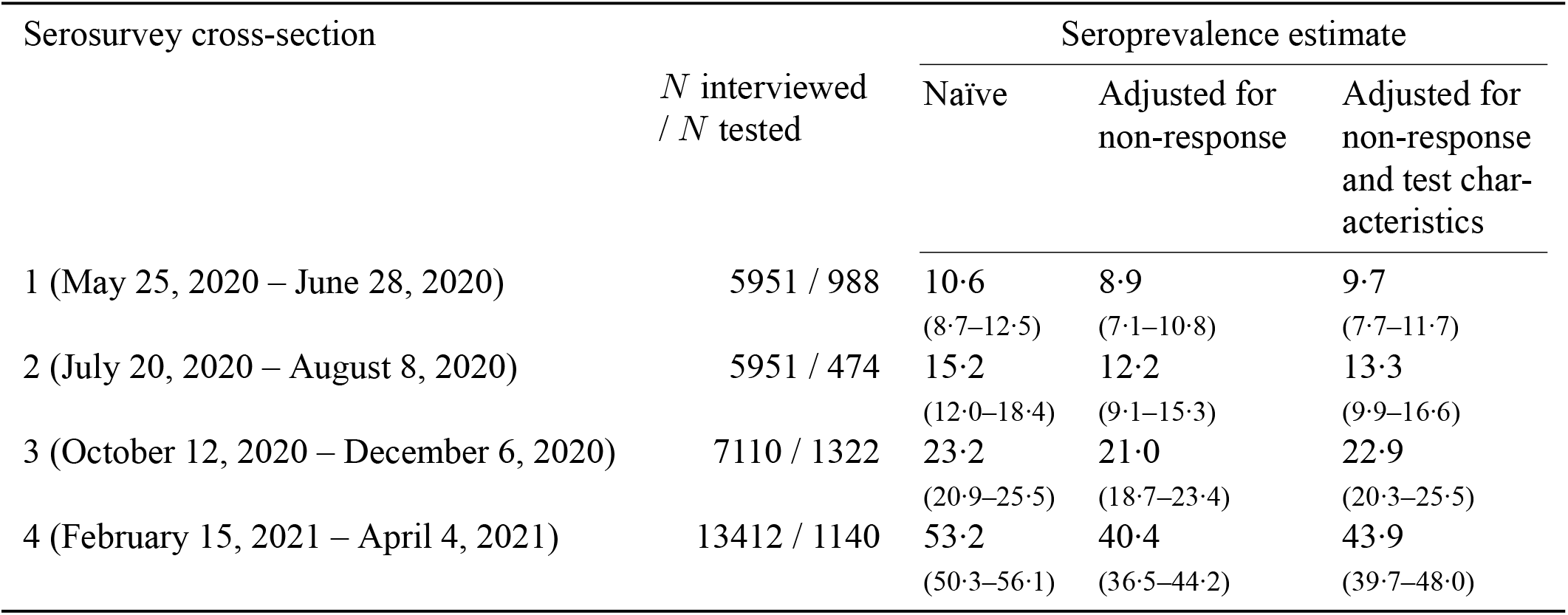
Seroprevalence by study cross-section, ELISA Coronapass

| Serosurvey cross-section | *N* interviewed<br>/ *N* tested | Seroprevalence estimate |  |  |
| --- | --- | --- | --- | --- |
|  |  | Naïve | Adjusted for<br>non-response | Adjusted for<br>non-response<br>and test char-<br>acteristics |
| 1 (May 25, 2020 – June 28, 2020) | 5951 / 988 | 10·6<br>(8·7–12·5) | 8·9<br>(7·1–10·8) | 9·7<br>(7·7–11·7) |
| 2 (July 20, 2020 – August 8, 2020) | 5951 / 474 | 15·2<br>(12·0–18·4) | 12·2<br>(9·1–15·3) | 13·3<br>(9·9–16·6) |
| 3 (October 12, 2020 – December 6, 2020) | 7110 / 1322 | 23·2<br>(20·9–25·5) | 21·0<br>(18·7–23·4) | 22·9<br>(20·3–25·5) |
| 4 (February 15, 2021 – April 4, 2021) | 13412 / 1140 | 53·2<br>(50·3–56·1) | 40·4<br>(36·5–44·2) | 43·9<br>(39·7–48·0) |

Seroprevalence estimates adjusted through raking weights were similar and are available in Supplementary Table S7. Sero-prevalence by different subgroups are reported in Supplementary Table S8. History of any symptoms, history of COVID-19 tests, and nonsmoking status were significant predictors for higher seroprevalence.

### Seroconversion results

The SARS-CoV-2 antibodies test results trajectories showed that most individuals remained seropositive with a maximum follow-up of 10 months (Figure 2). Among 177 participants who have reported at least one vaccine dose by the end of April, 2021, 92.7% (95% CI 87.9–95.7) were seropositive.

**Figure 2.**
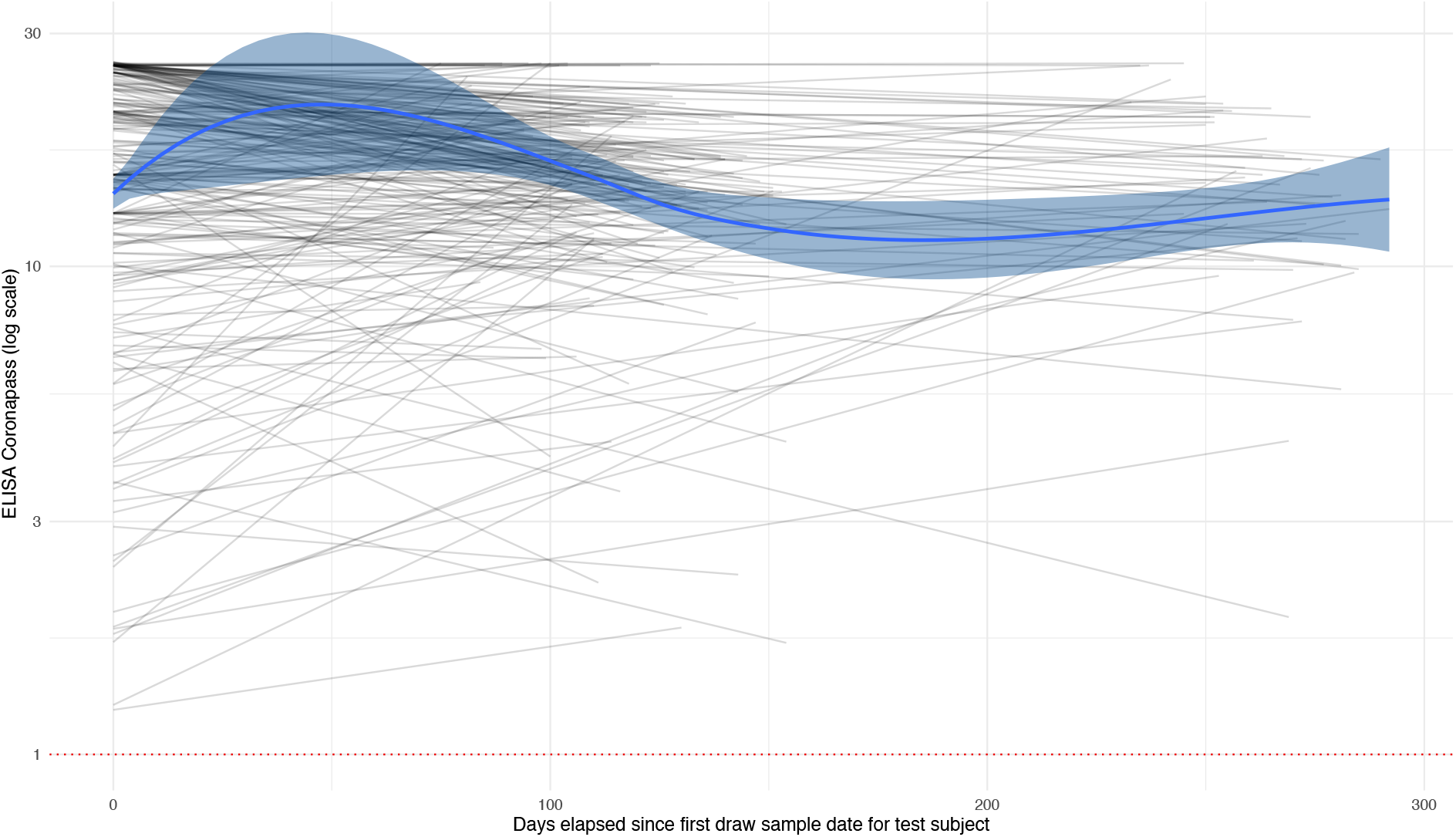
Trajectories of antibodies to SARS-CoV-2 (ELISA Coronapass). Grey lines are individual trajectories of study participants who tested positive at least once, excluding the 2020-07-20 – 2020-08-08 cross-section. Solid blue line is the loess smoother, blue areas report its 95% CI.

### Combining other sources of pandemic surveillance

The number of cases officially registered in the spring 2020 was much lower than in the autumn and the winter 2020–2021. Number of SARS-CoV-2 tests reached its maximum in the winter 2020–2021 in contrast to a relatively low number of tests reported in the spring 2020. Official case statistics contrast the number of hospitalisations, official deaths, and excess deaths reported in the spring 2020. The official number of cases, the number of hospitalisation and deaths from COVID-19 never reached zero between and after the pandemic waves. The number of COVID-19 deaths and excess deaths from all causes peaked in both periods and was in line with hospitalisation dynamics (Figure 3A).

**Figure 3.**
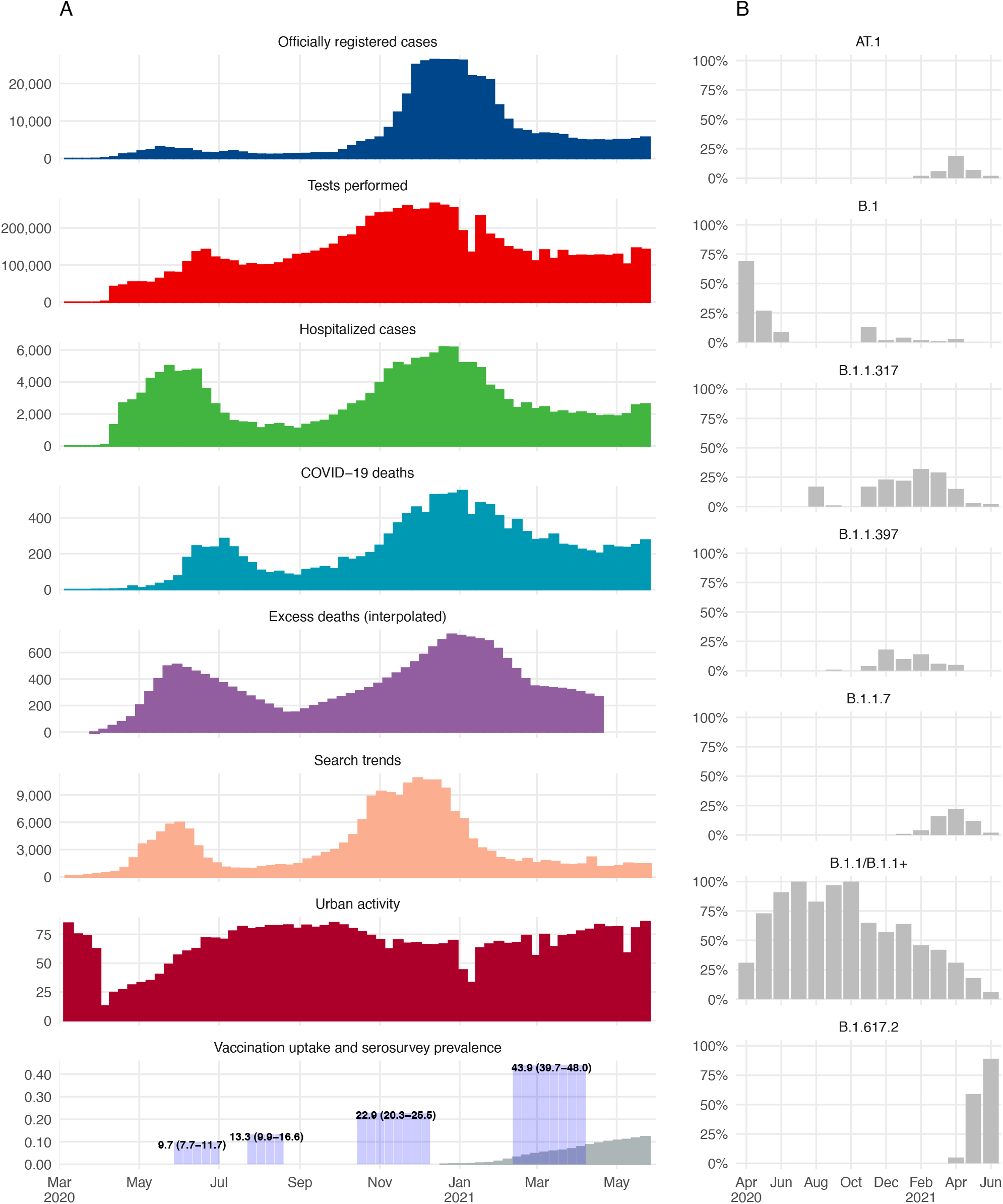
Combining available surveillance data to monitor the pandemic course in St. Petersburg during March-May 2020–2021:(A)Weekly data of officially registered cases, tests performed, hospitalised cases, COVID-19 deaths, interpolated excess deaths (from monthly data), search trends, urban activity, and vaccination uptake combined with seroprevalence estimates; (B) Monthly data on SARS-CoV-2 variants monitoring during April-June 2020–2021

Internet-based search terms trends were in line with pandemic dynamics. They reflected the changes in hospitalisation and death count better than the official case count, especially during the spring wave (Figure 3A). In addition, urban activity trends showed an apparent response to the first spring wave, somewhat less evident response during the second winter wave, and return to pre-pandemic activity levels in the late spring of 2021.

The SARS-CoV-2 circulating lineages diversity in 2020 was low. All samples from this period were attributed to the B.1 lineage and its sublineages. By autumn 2020 the number of PANGO lineages gradually increased with two Russian endemic — the B.1.397 and B.1.317. The Alpha VOC (B.1.1.7) was first detected in February 2021. The number of B.1.1.7 cases did not increase steeply but showed a gradual increase by April 2021. In February 2021, another lineage — AT.1, that has probably emerged in St. Petersburg was detected. The AT.1 was spreading rather quickly till April 2021, when B.1.617.2 (the Delta VOC) was first detected and substituted all other lineages by June 2021 (See Figure 3B).

### Infection fatality ratio

Using excess deaths data, the IFR was equal to 1.04 (95% CI 0.80–1.31) for the adult population for the whole pandemic period. IFR based on the official COVID-19 deaths counts was lower and amounted to 0.43% (95% CI 0.11–0.82). When we considered the entire population of the city rather than the adult population for IFR, we obtained the estimate of 0.86% (95% CI 0.66–1.08) based on the excess deaths data. Full results for IFR are reported in Supplementary Table S2. There was a clear upward trend in IFR by age. IFR was higher in men in all age groups.

## Discussion

Our study is the first comprehensive attempt to characterise the pandemic dynamics in the fourth largest European metropolitan area. We used all available sources for surveillance, including population-based seroprevalence study, the monitoring of SARS-CoV-2 VOCs, data on registered cases and deaths, relevant search term trends and city activity. Combining this data provides an overall global picture how the pandemic evolved through 2020 and 2021 in St. Petersburg. In April 2021, approximately one year after COVID-19, we estimated that about 45% contracted the SARS-CoV-2 infection in St. Petersburg, roughly 2.2 mln residents. Together with more than 10% vaccination uptake to that moment, less than 45% susceptibles were there in the population. Nevertheless, it was enough to avoid a new pandemic wave in the absence of mitigation measures till the spread of the Delta VOC (B.1.617.2) at the end of May 2021.

The first year of the COVID-19 pandemic in St. Petersburg can be characterised by two waves of similar intensity but different lengths. In the spring 2020, the first pandemic wave resulted in unprecedented mitigation measures and population reaction that helped flatten the pandemic curve and preserve the healthcare system functionality. As a result, the daily registered number of cases have plateaued in summer. It helped reorganise the hospital capacities in St. Petersburg and prepare for the subsequent increase in the case count. The hospitals had to experience the entire load in autumn and winter, although many additional beds were allocated to COVID-19 patients. The number of daily cases plateaued during the winter holidays went down to 700–900 officially registered cases per day in the spring 2020. The halt of most mitigation measures has not resulted in the subsequent pandemic wave until the Delta VOC started to spread rapidly in May 2021. The new summer 2021 pandemic wave is yet to be analysed.

Possibly the number of individuals who already have antibodies to SARS-CoV-2, which was reported to be a strong protection marker from reinfection [21] combined with mitigation measures still in place in winter, has played its role in the pandemic dynamics in 2021 in St. Petersburg. In our study, we did not see any seroreversion events with a maximum follow-up of ten months, which is in line with some other studies [22]. Population-based vaccination was introduced in St. Petersburg in early 2021 and progressed slowly but involved primarily individuals who have not contracted the disease. Therefore, the sum of individuals seropositive after infection and the vaccinated individuals can approximate the number of protected individuals, yielding around 50–55% individuals with antibodies to SARS-CoV-2 by the end of April 2021. However, this approximation may not be valid in the future as more and more individuals who contracted the disease proceed to vaccination.

One of the surprising findings, which other studies reproduce [23, 24], is an association between seropositivity and smoking status. Seroprevalence was lower for smokers. That association was evident for both population samples in our study. Our study, however, does not answer the question, whether smokers are less likely to be infected or to develop less durable protection against infection [25], which is more likely given higher IFR in men who smoke more often in Russia.

Internet search term trends were quite reliably reflecting the pandemic’s progress and predicted the increase in the number of hospitalisation and deaths for both waves in St. Petersburg. However, the Internet search term trends to monitor pandemics should be considered with caution [16]. This convenient surveillance option is compelling only in settings where the web-based search for medical conditions and symptoms is available and popular. Another critical limitation of the Internet search term trends lies in the spectrum of symptoms related to the disease of interest. For example, loss of smell is quite a distinct feature of SARS-CoV-2 infection. If the clinical manifestation of the infection caused by the new strains differ, surveillance strategies using search term trends should also change.

More than 20,000 excess deaths have already been reported in St. Petersburg during the pandemic year [11]. The results of our seroprevalence study combined with the data on excess mortality give the IFR equal to 0.86% for the entire population, which is in line with other estimates across Europe [7, 24]. The IFR based on serological study results and excess mortality was stable for all four surveys. However, the official COVID-19 death count provided lower IFRs, which were not stable and was even lower during the pandemic waves. Thus, it seems that the number of deaths during the both waves was unprecedented for St. Petersburg to timely provide official data collection and cause of death specifications in mortality records.

We continue to monitor the pandemic in St. Petersburg using all available sources and plan to run the following survey to estimate the number of individuals with antibodies to SARS-CoV-2 after the summer wave. In addition, we aim to detect the herd immunity threshold in St. Petersburg if any exists given the Delta VOC basic reproductive number and diminished vaccine effectiveness [12].

Several possible limitations of our serological survey may require further explanation. Small sample size and high non-response rate compared to the number of phone numbers generated pose a challenge in two cases. First, when the obtained sample is small enough to make the study underpowered. Our sample size calculations show that under the 50% hypothetical prevalence scenario, our sampling error does not exceed 3% [8]. Second, a high non-response rate is a problem when there is an unaccounted selection on observables or unobservables into the tested subsample. Under our study design, we observe a rich set of characteristics of individuals to account for non-response.

In conclusion, our study provided an overall description of SARS-CoV-2 pandemic progression in the fourth largest European city — St. Petersburg, Russia, using all available surveillance sources, including a population-based serological study to assess the prevalence of antibodies to SARS-CoV-2. More than a half of the city’s population had antibodies to the new coronavirus by April 2021, most of them due to prior infection. That was enough to control the SARS-CoV-2 without mitigation measure only until the Delta VOC universal spread. When compared against the number of overall excess deaths, our seroprevalence estimates align with the IFR of 0.86%. Furthermore, the combination of different surveillance sources, including internet search term trends, provide a clear picture of the course of the SARS-CoV-2 pandemic in St. Petersburg.

## Data Availability

All analyses were conducted in R, study data and code is available online (https://github.com/eusporg/spb_covid_study20).

## Authors’ contributions

Anton Barchuk, Dmitriy Skougarevskiy, Kirill Titaev, Lubov Barabanova, Daniil Shirokov, Daria Danilenko, and Dmitry Lioznov conceived the study. Anton Barchuk, Dmitriy Skougarevskiy, Daniil Shirokov, and Daria Danilenko coordinated the study. Anton Barchuk, Dmitriy Skougarevskiy, Alexei Kouprianov, Daria Danilenko, Andrey Komissarov, Rustam Tursun-zade, Konstantin Blagodatskikh, and Olga Dudkina drafted the first version of the manuscript. Kirill Titaev, Mariia Sergeeva, Varvara Tychkova, Alena Zheltukhina, Dmitry Lioznov, Artur Isaev, Ekaterina Pomerantseva, Svetlana Zhikrivetskaya, Konstantin Blagodatskikh, Yana Sofronova, Lubov Barabanova, and Daniil Shirokov contributed to drafting sections of the manuscript. Dmitriy Skougarevskiy, Anton Barchuk, Alexei Kouprianov, Andrey Komissarov, and Daria Danilenko did data analyses. Mariia Sergeeva, Varvara Tychkova, Andrey Komissarov, Alena Zheltukhina, Dmitry Lioznov, Artur Isaev, Ekaterina Pomerantseva, Yana Sofronova, Svetlana Zhikrivetskaya, and Konstantin Blagodatskikh did lab analyses. All authors participated in the study design, helped to draft the manuscript, contributed to the interpretation of data and read and approved the final manuscript.

## Funding

Polymetal International plc funded the serological study. The main funder had no role in study design, data collection, data analysis, data interpretation, writing of the report or decision to submit the publication. The European University at St. Petersburg, clinic “Scandinavia”, Smorodintsev Research Institute of Influenza and Genetico had access to the study data. The European University at St. Petersburg had final responsibility for the decision to submit for publication. Part of this study performed at Smorodintsev Research Institute of Influenza was funded by the Russian Ministry of Science and Higher Education as part of the Worldclass Research Center program: Advanced Digital Technologies (contract No. 075152020904, dated 16.11.2020).

## Declaration of interests

Anton Barchuk reports personal fees from AstraZeneca, MSD, and Biocad outside the submitted work. Other authors have no conflict of interest to declare.

## Acknowledgements

We acknowledge personal support from Vitaly Nesis (Chief Executive Officer, Polymetal International, plc). We thank Alla Samoletova (European University at St. Petersburg) for administrative support and management of the study, Yulia Stepantsova (Chursina) for coordinating phone-based interviews, Lizaveta Dubovik, and Irina Shubina for the science communication. We thank the interviewers, nurses, general practitioners, and the Clinic “Scandinavia” personnel. We also thank all study participants.

## Supplementary materials

### Data appendix

The federal government and St. Petersburg city government made most of the detailed statistics regarding COVID-19 available. However the data are scattered across different sources. The number of daily new cases was obtained from the official federal government website (`stopcoronavirus.rf`, https://xn--80aesfpebagmfblc0a.xn--plai/). The daily reports of new cases were also provided by the St. Petersburg city government and the city Health Committee. While the official website provided a somewhat smoothed pandemic curve, the St. Petersburg city government and Health Committee data looked closer to sources from other countries, with seasonal fluctuations on weekends and holidays. However, the city data are available only from early December 2020. The number of COVID-19 deaths was obtained from the official data from `stopcoronavirus.rf`. The number of excess deaths was obtained from Kobak (2020), the calculation was based on monthly data from the Federal State Statistics Service of Russia on deaths from any causes [11]. This data is constructed by subtracting the linear trend in monthly deaths over 2015–19 from the monthly deaths data in 2020–21 and is available online https://github.com/dkobak/excess-mortality. We obtained the number of tests to detect SARS-CoV-2 from the official St. Petersburg city government Telegram channel https://t.me/koronavirusspb. The number of hospital admissions are extracted from the St. Petersburg city government and Health Committee reports. The ongoing surveillance for SARS-CoV-2 VOCs in St. Petersburg is carried out by the Smorodintsev Research Institute of Influenza and is described in detail elsewhere [18].

### Statistical appendix: estimating the IR and IFR with the Bayesian evidence synthesis model

#### Observables

We conduct *K* cross-sections of serosurvey of adult population of St. Petersburg, Russia. In each cross-section *k* = 1,…,*K* we randomly select *T*_*k*_ individuals to get tested out of *P*_*k*_ individuals at risk of infection in the city. Out of those tested we identify *CC*_*k*_ seropositive individuals with confirmed cases of SARS-CoV-2. In each cross-section *k* we also observe the cumulative number of deaths *D*_*k*_ attributed to COVID-19 since the pandemic onset in the city.

#### Latent variables

We need to make inference on following variables:

- *C*_*k*_ — the cumulative total number of infected individuals by wave *k*,
- *IR*_*k*_ — the true infection rate (proportion of population which has been infected by cross-section *k*), which is the expected value E[*Ck/Pk* ],
- *IFR*_*k*_ — the true underlying infection fatality rate, which is the expected value E[*Dk/Ck* |*C*_*k*_].

To estimate the IFR across the study cross-sections we closely follow [19] who proposed a simple framework for Bayesian evidence synthesis.

#### Distributional and modeling assumptions

We make the following assumptions on the distribution of the latent variables:

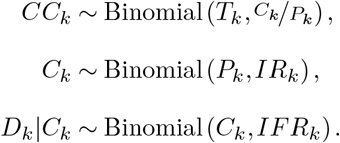

Following [19], to improve the MCMC mixing we replace the assumption for *CC*_*k*_ with *CC*_*k*_ *∼* Binomial(*T*_*k*_,*IFR*_*k*_). Then we can replace the conditional assumption *D*_*k*_|*C*_*k*_ with the unconditional

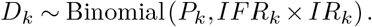

Next we assume that per-cross-section *IFR*_*k*_ and *IR*_*k*_ are distributed according to a random effects model:

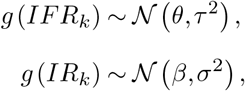

where *g* (*•*) *≡* log(*-* log(1 *-•*)) is the complimentary log-log link function, *θ* is the mean clog-log-transformed infection fatality rate across the study cross-sections, *τ* reflects the variability in between-cross-section IFR estimates, *β* is the mean clog-log-transformed infection rate across the cross-sections, *σ* captures the variability in between-cross-section IR estimates.

#### Prior elicitation

Following [19, 26] we consider two sets of priors on model parameters:

- Weakly informative priors: *g*^*-*1^ (*θ*) *∼* Beta(0.3, 30), *g*^*-*1^ (*β*) *∼* Beta(1, 30), *σ ∼* half-*𝒩* (0, 10), *τ ∼* half-*𝒩* (0, 10);
- Non-informative flat priors: *g*^*-*1^ (*θ*) *∼* Uniform(0, 1), *g*^*-*1^ (*β*) *∼* Uniform(0, 1), *σ ∼* half-*𝒩* (0, 100), *τ ∼* half-*𝒩* (0, 100).

#### Estimation and inference

The model is fit with JAGS [27] with 5 independent chains, each with 2 million draws (20% burn-in, thinning of 100). We then report the median estimates of per-cross-section *IFR*_*k*_ and *IR*_*k*_ and their 95% highest probability density credible intervals [28].

#### Data

To fit the model we need to acknowledge multiple data constraints. For *P*_*k*_ we assume that the entire adult (*≥* 18 years old) population of the city is at risk of infection (in a sensitivity analysis we consider the entire city population instead). For cross-sections one and two we take the adult city population count as of January 1, 2020 from the Federal State Statistics Service of Russia*, 4 451 025 individuals. The data on the adult population as of January 1, 2021 is not available at the time of writing of this paper. However, the official data on the total city population is available^†^ and amounts to 5 384 342 people as of January 1, 2021 and 5 398 064 as of January 1, 2020. We assume that the adult population followed the same trend as the total population (a -0.25% decline) in 2020 and assume *P*_*k*_ = 4 451 025 *×* (1 *-* 0 *·* 0025) = 4 439 897 individuals for cross-sections three and four.

We do not take the values *T*_*k*_ and *CC*_*k*_ directly from the per-cross-section test data. To arrive at the seroprevalence estimate in our study we adjusted those naïve figures for test performance and non-response bias. Instead of using the raw counts we invert the reported 95% CI for the seroprevalence estimate for cross-section *k*. Using a beta prior on the probability of success for a binomial distribution, we can determine a two-sided confidence interval from a beta posterior for any given *T*_*k*_ and *CC*_*k*_. We define the values of *T*_*k*_ and *CC*_*k*_ that correspond to the reported seroprevalence 95% CI for ELISA Coronapass from Table 1 adjusted for non-response and test characteristics as the effective 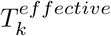 and 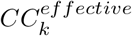 and use those values in the model. Such “inverting uncertainty intervals” approach of [19] allows us to easily incorporate our seroprevalence adjustments coming from a frequentist unnivariate imputation model into a Bayesian evidence synthesis model.

When it comes to the cumulative number of deaths *D*_*k*_ by cross-section *k* an obvious question is what date to use to compute this figure for each study cross-section. [29] suggest compute the total number of deaths up until seven days after the cross-section mid-point. [19] propose to treat *D*_*k*_ as an interval censored variable where we do not know its true value but observe its lower and upper bounds 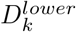 and 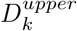 for each cross-section. The authors define 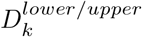 as the total number of deaths from the pandemic onset until 14 days after the start/the end of the cross-section *k*, respectively. We adopt this approach as it allows for uncertainty in the actual death counts.

Another concern is reliability of the reported deaths data. We use two sources for 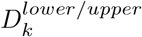. The first is the official national government website (`stopcorovirus.rf`) that provides daily data on COVID-related deaths in St. Petersburg. The second is excess deaths estimation based on monthly data from the Federal State Statistics Service of Russia [11]. We find it valuable to compute the IFR and IR using the data from both sources given the voiced concerns about under-reporting of COVID-related deaths in the country. For monthly excess deaths data we consider the cumulative excess deaths from January 1, 2020 to the month of the cross-section start to define 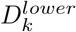 and the cumulative excess deaths from January 1, 2020 to the month of the cross-section end to define 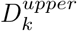. All the variables used in the estimation are reported in Table S1.

**Table S1.**
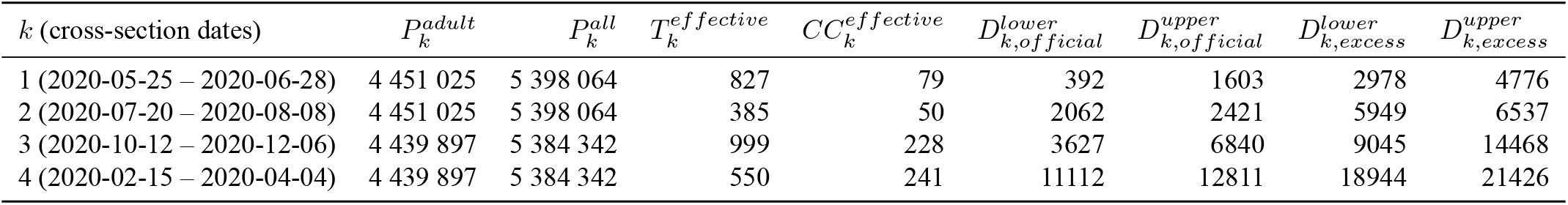
Data used for IR/IFR estimation in the Bayesian evidence synthesis model

## Results

The per-cross-section estimates of IR/IFR under different priors and death intervals are reported in Table S2.

### Per-age and sex IR and IFR

Our approach can be easily applied to another problem. Suppose now that *k* indexes sex and age groups within one serosurvey cross-section. Then we can use the same logic to estimate IR and IFR for each age group-sex combinations.

We predict ELISA Coronapass-based seroprevalence within each sex and age group combination from our baseline univariate model for cross-section 4 where we use an interaction between individual age group and sex instead of treating them as linearly separable variables (as reported in Table S8) and define more fine-grained age groups. Then we invert the estimated CI for the seroprevalence to compute the 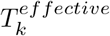 and 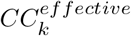 (see Table S3). For per-group population *P*_*k*_ we use data as of the beginning of 2020 since no data for 2021 is available yet.

**Table S2.**
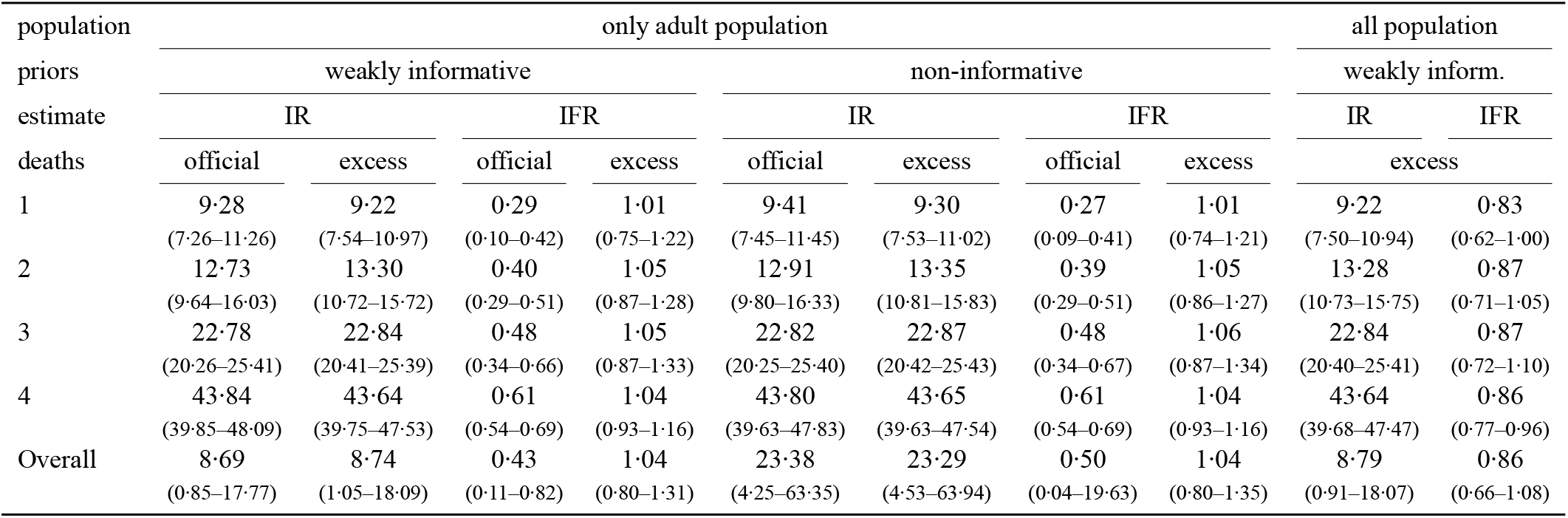
Estimated IR/IFR across the study cross-sections from the Bayesian evidence synthesis model

When it comes to 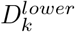 and 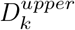 we need to acknowledge that, to the best of our knowledge, no official data on deaths from COVID-19 disaggregated by age and sex exists. For this reason, we rely on excess deaths data estimation. We gather official yearly data on deaths in 2016–19 by age group and sex and quarterly data on deaths in 2020–21 to compute our 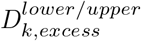. We used quarterly data of age and sex-specific number of deaths and population from the Federal State Statistics Service of Russia. First, we combined the number of deaths from all causes from the second, third, and fourth quarter of 2020, and the first quarter of 2021 (the pandemic year). We treated the pandemic year as a calendar year, as it captures all seasonal trends, and includes all periods when excess deaths due to COVID-19 are expected. The first case of SARS-CoV-2 infection was registered in Saint Petersburg in Russia on March 5, 2020, and it is not likely that the first quarter of 2020 contributed to excess mortality due to COVID-19. We estimated expected deaths by using a Poisson model that accounts for annual temporal trends within each age and sex-specific group with an offset that accounts for the population size in each group. For each age and sex-specific group, the model used mortality data for 2016-19 to estimate the expected number of deaths in each group for 2020. Then the predicted lower and upper bound for expected death count in 2020 was used to estimate the number of excess death in the pandemic year. The cumulative number of deaths across all age and sex-specific groups combined (the lower bound was 18 631 and the upper was 22 289) was in line with the excess deaths estimation based on monthly data from the Federal State Statistics Service of Russia (the lower bound was 18 944 and the upper 21 426) [11].

The estimated IR and IFR for each age and sex-specific group combination are in Table S4.

**Table S3.**
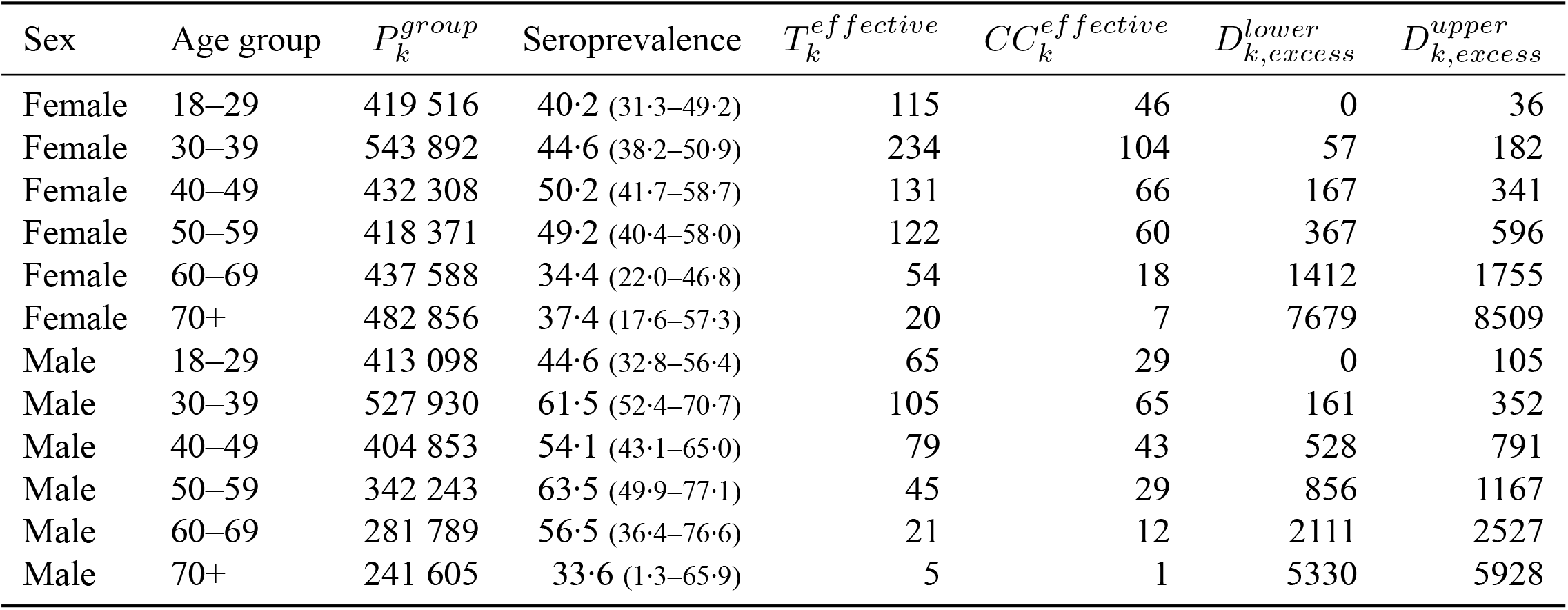
Data used for IR/IFR estimation for cross-section 4 (ELISA Coronapass) in the Bayesian evidence synthesis model by age group and sex

**Table S4.**
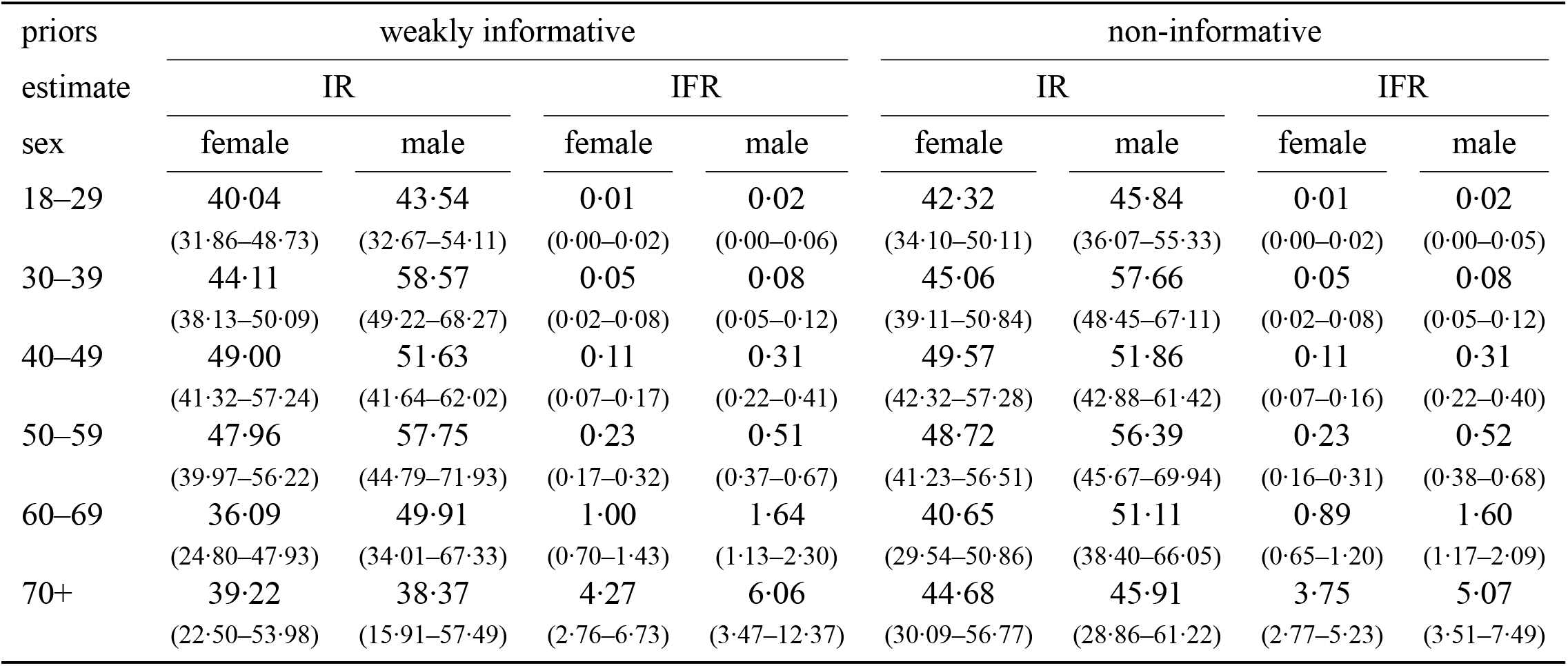
Estimated IR/IFR across the age and sex groups from the Bayesian evidence synthesis model, cross-section 4, ELISA Coronapass

**Table S5.**
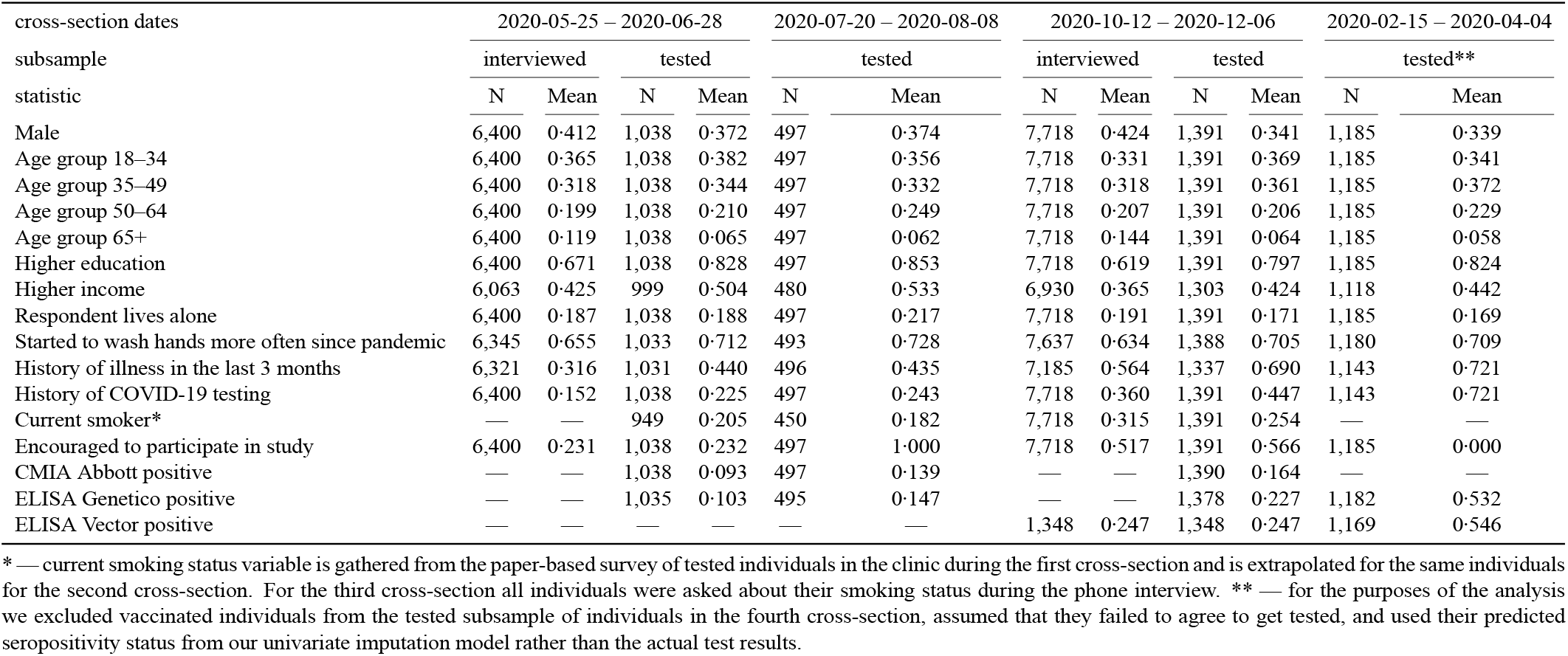
Summary statistics across study cross-sections

**Table S6.**
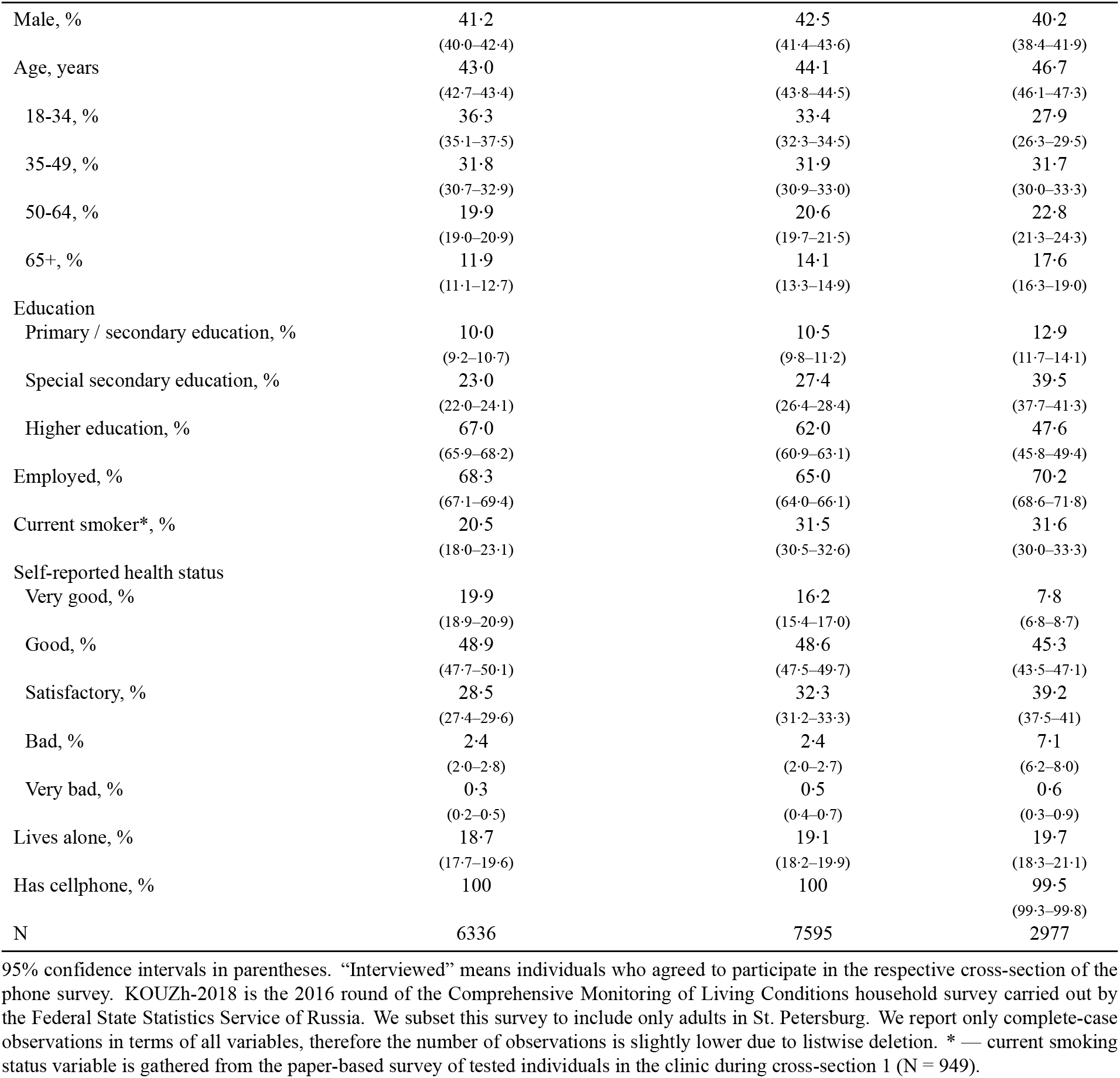
Representativeness of the survey across study cross-sections

**Figure S1.**
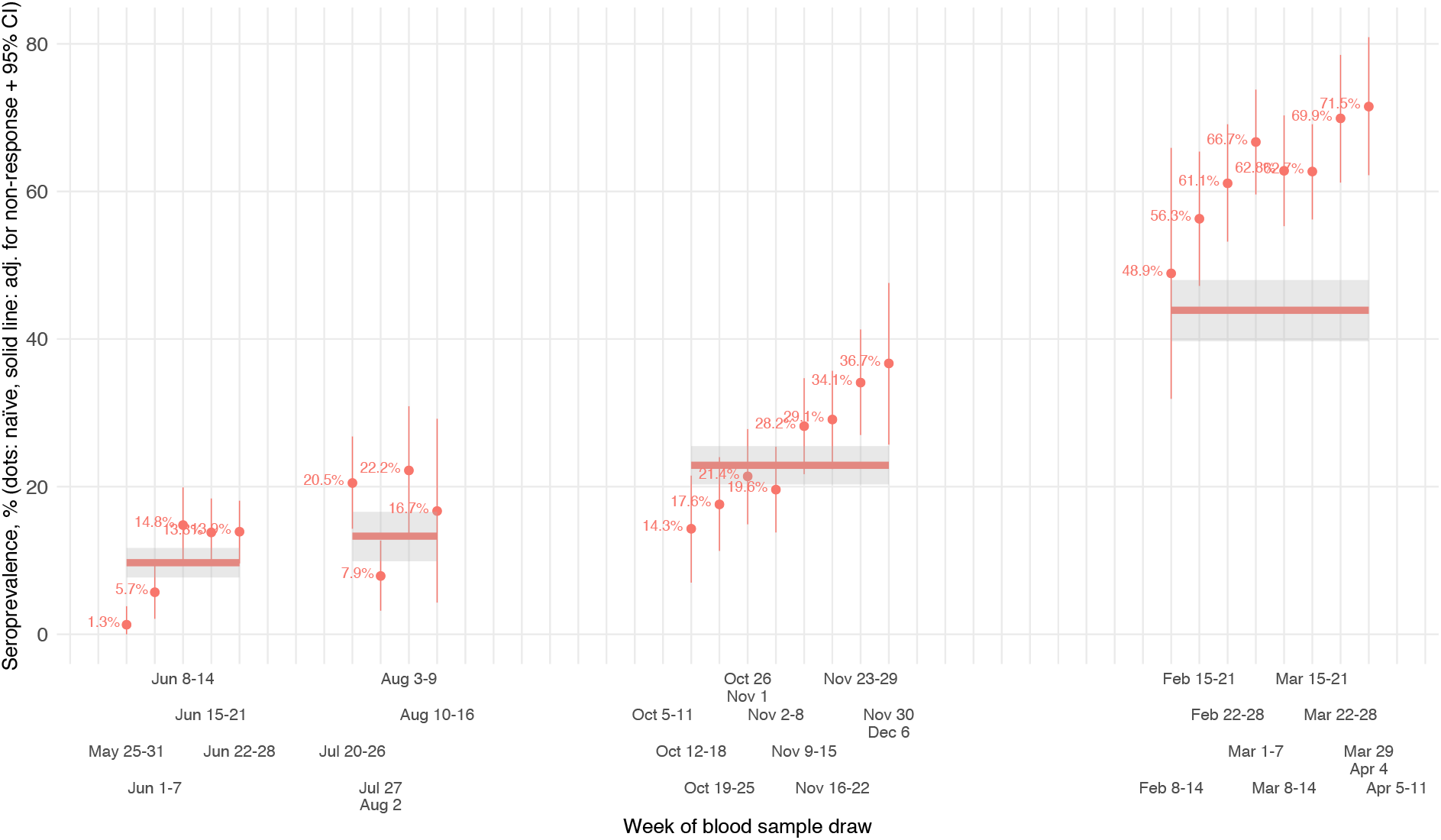
Naïve and adjusted seroprevalence by study cross-section and week (ELISA Coronapass)

**Table S7.**
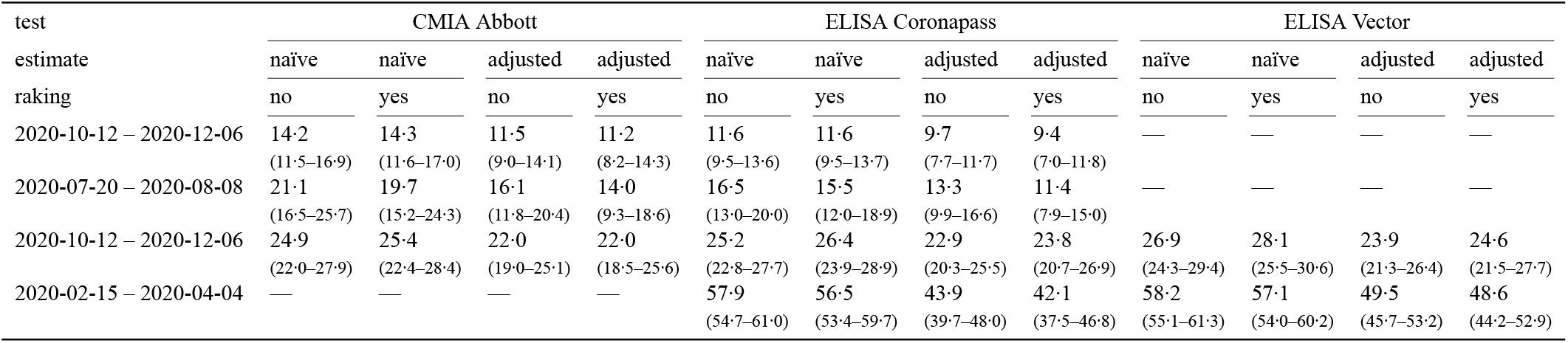
Seroprevalence by cross-section: naïve adjusted for non-response bias or adjusted for non-response and test performance

**Table S8.**
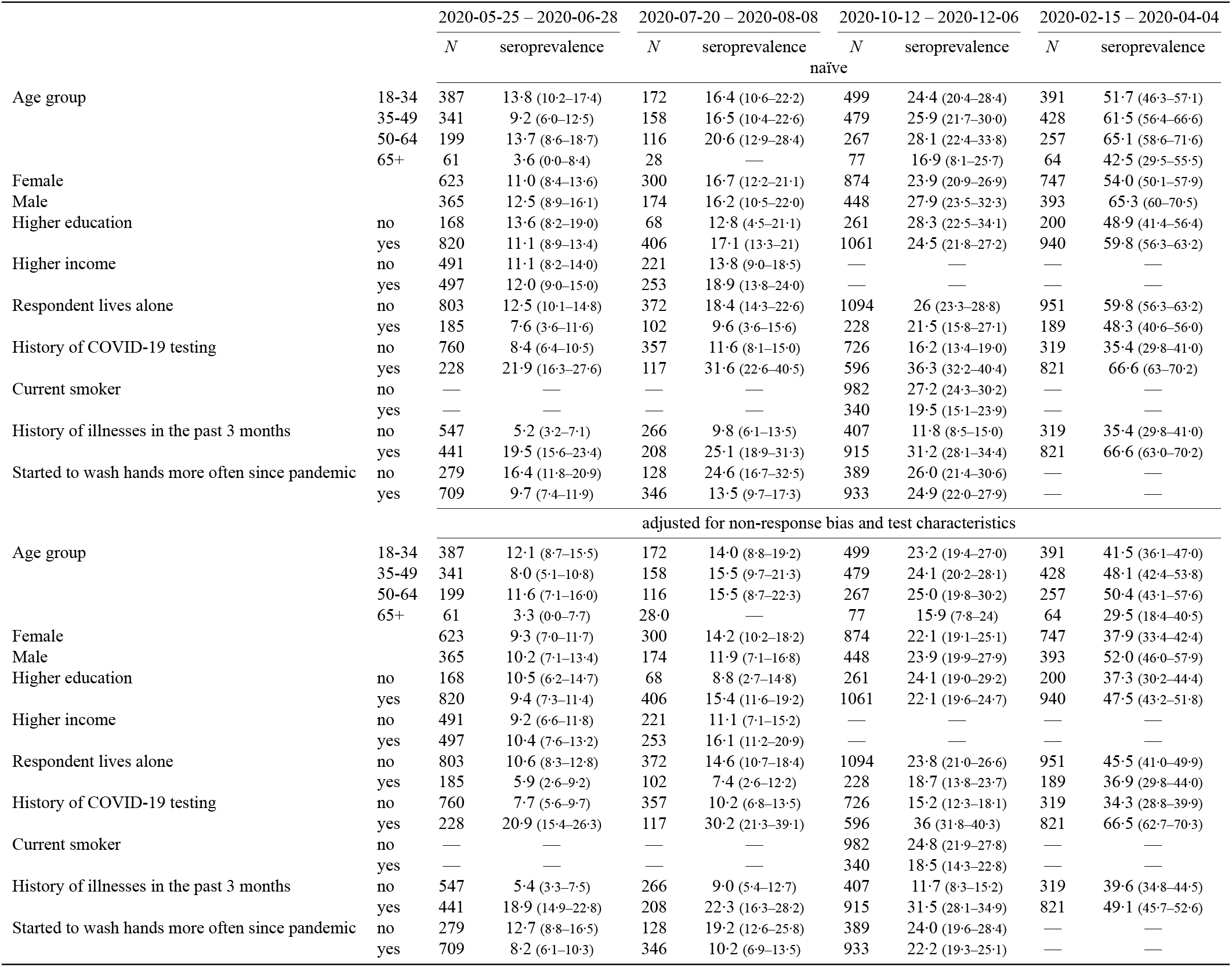
Seroprevalence by subgroup, ELISA Coronapass

## Footnotes

* https://gks.ru/bgd/regl/b20_111/Main.htm

† https://petrostat.gks.ru/folder/27595

## Notes

### Clinical Trial

ISRCTN11060415

### Clinical Protocols

https://eusp.org/sites/default/files/inline-files/EU_SG-Russian-Covid-Serosurvey-Protocol-CDRU-001_en.pdf

### Author Declarations

The Research Planning Board approved the study of the European University at St. Petersburg (on May 20, 2020) and the Ethics Committee of the Clinic Scandinavia (on May 26, 2020). All research was performed following the relevant guidelines and regulations. Informed consent was obtained from all participants of the study. The study was registered with the following identifiers: Clinicaltrials.gov (NCT04406038, submitted on May 26, 2020, date of registration - May 28, 2020) and ISRCTN registry (ISRCTN11060415, submitted on May 26, 2020, date of registration - May 28, 2020). Official statistics, VOCs monitoring data, search terms trends, and mobility trends were obtained from open sources as aggregated data. Analysis based on open-source aggregated data does not require additional ethical permission in Russia.

