## Supplementary material for "COVID-19 pandemic in Saint Petersburg, Russia: combining surveillance and population-based serological study data in May, 2020–April, 2021"

**Latent variables** We need to make inference on following variables:

- $C_k$ — the cumulative total number of infected individuals by wave $k$,
- $IR_k$ — the true infection rate (proportion of population which has been infected by cross-section $k$), which is the expected value $\mathrm{E}\left[{}^{C_k}/_{P_k}\right]$,
- $IFR_k$ — the true underlying infection fatality rate, which is the expected value $\mathrm{E}\left[{}^{D_k}/_{C_k} | C_k\right]$.

To estimate the IFR across the study cross-sections we closely follow [19] who proposed a simple framework for Bayesian evidence synthesis.

**Distributional and modeling assumptions** We make the following assumptions on the distribution of the latent variables:

$$CC_k \sim \text{Binomial}\left(T_k, {}^{C_k}/_{P_k}\right),$$
$$C_k \sim \text{Binomial}\left(P_k, IR_k\right),$$
$$D_k|C_k \sim \text{Binomial}\left(C_k, IFR_k\right).$$

Following [19], to improve the MCMC mixing we replace the assumption for $CC_k$ with $CC_k \sim \text{Binomial}\left(T_k, IFR_k\right)$. Then we can replace the conditional assumption $D_k|C_k$ with the unconditional

$$D_k \sim \text{Binomial}\left(P_k, IFR_k \times IR_k\right).$$

Next we assume that per-cross-section $IFR_k$ and $IR_k$ are distributed according to a random effects model:

$$g\left(IFR_k\right) \sim \mathcal{N}\left(\theta, \tau^2\right),$$
$$g\left(IR_k\right) \sim \mathcal{N}\left(\beta, \sigma^2\right),$$

where $g\left(\bullet\right) \equiv \log\left(-\log\left(1-\bullet\right)\right)$ is the complimentary log-log link function, $\theta$ is the mean clog-log-transformed infection fatality rate across the study cross-sections, $\tau$ reflects the variability in between-cross-section IFR estimates, $\beta$ is the mean clog-log-transformed infection rate across the cross-sections, $\sigma$ captures the variability in between-cross-section IR estimates.

**Prior elicitation** Following [19, 26] we consider two sets of priors on model parameters:

- Weakly informative priors: $g^{-1}\left(\theta\right) \sim \text{Beta}\left(0.3, 30\right)$, $g^{-1}\left(\beta\right) \sim \text{Beta}\left(1, 30\right)$, $\sigma \sim \text{half-}\mathcal{N}\left(0, 10\right)$, $\tau \sim \text{half-}\mathcal{N}\left(0, 10\right)$;
- Non-informative flat priors: $g^{-1}\left(\theta\right) \sim \text{Uniform}\left(0, 1\right)$, $g^{-1}\left(\beta\right) \sim \text{Uniform}\left(0, 1\right)$, $\sigma \sim \text{half-}\mathcal{N}\left(0, 100\right)$, $\tau \sim \text{half-}\mathcal{N}\left(0, 100\right)$.

**Estimation and inference** The model is fit with JAGS [27] with 5 independent chains, each with 2 million draws (20% burn-in, thinning of 100). We then report the median estimates of per-cross-section $IFR_k$ and $IR_k$ and their 95% highest probability density credible intervals [28].

**Data** To fit the model we need to acknowledge multiple data constraints. For $P_k$ we assume that the entire adult ($\geq 18$ years old) population of the city is at risk of infection (in a sensitivity analysis we consider the entire city population instead). For cross-sections one and two we take the adult city population count as of January 1, 2020 from the Federal State Statistics Service of Russia*, 4 451 025 individuals. The data on the adult population as of January 1, 2021 is not available at the time of writing of this paper. However, the official data on the total city population is available† and amounts to 5 384 342 people as of January 1,

*https://gks.ru/bgd/regl/b20_111/Main.htm
†https://petrostat.gks.ru/folder/27595

2021 and 5 398 064 as of January 1, 2020. We assume that the adult population followed the same trend as the total population (a -0.25% decline) in 2020 and assume $P_k = 4\ 451\ 025 \times (1 - 0 \cdot 0025) = 4\ 439\ 897$ individuals for cross-sections three and four.

**Table S1.** Data used for IR/IFR estimation in the Bayesian evidence synthesis model

| $k$ (cross-section dates) | $P_k^{adult}$ | $P_k^{all}$ | $T_k^{effective}$ | $CC_k^{effective}$ | $D_{k,official}^{lower}$ | $D_{k,official}^{upper}$ | $D_{k,excess}^{lower}$ | $D_{k,excess}^{upper}$ |
|---|---|---|---|---|---|---|---|---|
| 1 (2020-05-25 – 2020-06-28) | 4 451 025 | 5 398 064 | 827 | 79 | 392 | 1603 | 2978 | 4776 |
| 2 (2020-07-20 – 2020-08-08) | 4 451 025 | 5 398 064 | 385 | 50 | 2062 | 2421 | 5949 | 6537 |
| 3 (2020-10-12 – 2020-12-06) | 4 439 897 | 5 384 342 | 999 | 228 | 3627 | 6840 | 9045 | 14468 |
| 4 (2020-02-15 – 2020-04-04) | 4 439 897 | 5 384 342 | 550 | 241 | 11112 | 12811 | 18944 | 21426 |

**Results** The per-cross-section estimates of IR/IFR under different priors and death intervals are reported in Table S2.

**Per-age and sex IR and IFR** Our approach can be easily applied to another problem. Suppose now that $k$ indexes sex and age groups within one serosurvey cross-section. Then we can use the same logic to estimate IR and IFR for each age group-sex combinations.

We predict ELISA Coronapass-based seroprevalence within each sex and age group combination from our baseline univariate

**Table S2.** Estimated IR/IFR across the study cross-sections from the Bayesian evidence synthesis model

| population | only adult population | | | | | | | | all population | |
|---|---|---|---|---|---|---|---|---|---|---|
| priors | weakly informative | | | | non-informative | | | | weakly inform. | |
| estimate | IR | | IFR | | IR | | IFR | | IR | IFR |
| deaths | official | excess | official | excess | official | excess | official | excess | excess | |
| 1 | 9·28 (7·26–11·26) | 9·22 (7·54–10·97) | 0·29 (0·10–0·42) | 1·01 (0·75–1·22) | 9·41 (7·45–11·45) | 9·30 (7·53–11·02) | 0·27 (0·09–0·41) | 1·01 (0·74–1·21) | 9·22 (7·50–10·94) | 0·83 (0·62–1·00) |
| 2 | 12·73 (9·64–16·03) | 13·30 (10·72–15·72) | 0·40 (0·29–0·51) | 1·05 (0·87–1·28) | 12·91 (9·80–16·33) | 13·35 (10·81–15·83) | 0·39 (0·29–0·51) | 1·05 (0·86–1·27) | 13·28 (10·73–15·75) | 0·87 (0·71–1·05) |
| 3 | 22·78 (20·26–25·41) | 22·84 (20·41–25·39) | 0·48 (0·34–0·66) | 1·05 (0·87–1·33) | 22·82 (20·25–25·40) | 22·87 (20·42–25·43) | 0·48 (0·34–0·67) | 1·06 (0·87–1·34) | 22·84 (20·40–25·41) | 0·87 (0·72–1·10) |
| 4 | 43·84 (39·85–48·09) | 43·64 (39·75–47·53) | 0·61 (0·54–0·69) | 1·04 (0·93–1·16) | 43·80 (39·63–47·83) | 43·65 (39·63–47·54) | 0·61 (0·54–0·69) | 1·04 (0·93–1·16) | 43·64 (39·68–47·47) | 0·86 (0·77–0·96) |
| Overall | 8·69 (0·85–17·77) | 8·74 (1·05–18·09) | 0·43 (0·11–0·82) | 1·04 (0·80–1·31) | 23·38 (4·25–63·35) | 23·29 (4·53–63·94) | 0·50 (0·04–19·63) | 1·04 (0·80–1·35) | 8·79 (0·91–18·07) | 0·86 (0·66–1·08) |

The estimated IR and IFR for each age and sex-specific group combination are in Table S4.

**Table S3.** Data used for IR/IFR estimation for cross-section 4 (ELISA Coronapass) in the Bayesian evidence synthesis model by age group and sex

| Sex | Age group | $P_k^{group}$ | Seroprevalence | $T_k^{effective}$ | $CC_k^{effective}$ | $D_{k,excess}^{lower}$ | $D_{k,excess}^{upper}$ |
|---|---|---|---|---|---|---|---|
| Female | 18–29 | 419 516 | 40·2 (31·3–49·2) | 115 | 46 | 0 | 36 |
| Female | 30–39 | 543 892 | 44·6 (38·2–50·9) | 234 | 104 | 57 | 182 |
| Female | 40–49 | 432 308 | 50·2 (41·7–58·7) | 131 | 66 | 167 | 341 |
| Female | 50–59 | 418 371 | 49·2 (40·4–58·0) | 122 | 60 | 367 | 596 |
| Female | 60–69 | 437 588 | 34·4 (22·0–46·8) | 54 | 18 | 1412 | 1755 |
| Female | 70+ | 482 856 | 37·4 (17·6–57·3) | 20 | 7 | 7679 | 8509 |
| Male | 18–29 | 413 098 | 44·6 (32·8–56·4) | 65 | 29 | 0 | 105 |
| Male | 30–39 | 527 930 | 61·5 (52·4–70·7) | 105 | 65 | 161 | 352 |
| Male | 40–49 | 404 853 | 54·1 (43·1–65·0) | 79 | 43 | 528 | 791 |
| Male | 50–59 | 342 243 | 63·5 (49·9–77·1) | 45 | 29 | 856 | 1167 |
| Male | 60–69 | 281 789 | 56·5 (36·4–76·6) | 21 | 12 | 2111 | 2527 |
| Male | 70+ | 241 605 | 33·6 (1·3–65·9) | 5 | 1 | 5330 | 5928 |

**Table S4.** Estimated IR/IFR across the age and sex groups from the Bayesian evidence synthesis model, cross-section 4, ELISA Coronapass

| priors | weakly informative | | | | non-informative | | | |
|---|---|---|---|---|---|---|---|---|
| estimate | IR | | IFR | | IR | | IFR | |
| sex | female | male | female | male | female | male | female | male |
| 18–29 | 40·04 (31·86–48·73) | 43·54 (32·67–54·11) | 0·01 (0·00–0·02) | 0·02 (0·00–0·06) | 42·32 (34·10–50·11) | 45·84 (36·07–55·33) | 0·01 (0·00–0·02) | 0·02 (0·00–0·05) |
| 30–39 | 44·11 (38·13–50·09) | 58·57 (49·22–68·27) | 0·05 (0·02–0·08) | 0·08 (0·05–0·12) | 45·06 (39·11–50·84) | 57·66 (48·45–67·11) | 0·05 (0·02–0·08) | 0·08 (0·05–0·12) |
| 40–49 | 49·00 (41·32–57·24) | 51·63 (41·64–62·02) | 0·11 (0·07–0·17) | 0·31 (0·22–0·41) | 49·57 (42·32–57·28) | 51·86 (42·88–61·42) | 0·11 (0·07–0·16) | 0·31 (0·22–0·40) |
| 50–59 | 47·96 (39·97–56·22) | 57·75 (44·79–71·93) | 0·23 (0·17–0·32) | 0·51 (0·37–0·67) | 48·72 (41·23–56·51) | 56·39 (45·67–69·94) | 0·23 (0·16–0·31) | 0·52 (0·38–0·68) |
| 60–69 | 36·09 (24·80–47·93) | 49·91 (34·01–67·33) | 1·00 (0·70–1·43) | 1·64 (1·13–2·30) | 40·65 (29·54–50·86) | 51·11 (38·40–66·05) | 0·89 (0·65–1·20) | 1·60 (1·17–2·09) |
| 70+ | 39·22 (22·50–53·98) | 38·37 (15·91–57·49) | 4·27 (2·76–6·73) | 6·06 (3·47–12·37) | 44·68 (30·09–56·77) | 45·91 (28·86–61·22) | 3·75 (2·77–5·23) | 5·07 (3·51–7·49) |

### **Table S5.** Summary statistics across study cross-sections

| cross-section dates | 2020-05-25 – 2020-06-28 | | | | 2020-07-20 – 2020-08-08 | | 2020-10-12 – 2020-12-06 | | | | 2020-02-15 – 2020-04-04 | |
|---|---|---|---|---|---|---|---|---|---|---|---|---|
| subsample | interviewed | | tested | | tested | | interviewed | | tested | | tested** | |
| statistic | N | Mean | N | Mean | N | Mean | N | Mean | N | Mean | N | Mean |
| Male | 6,400 | 0·412 | 1,038 | 0·372 | 497 | 0·374 | 7,718 | 0·424 | 1,391 | 0·341 | 1,185 | 0·339 |
| Age group 18–34 | 6,400 | 0·365 | 1,038 | 0·382 | 497 | 0·356 | 7,718 | 0·331 | 1,391 | 0·369 | 1,185 | 0·341 |
| Age group 35–49 | 6,400 | 0·318 | 1,038 | 0·344 | 497 | 0·332 | 7,718 | 0·318 | 1,391 | 0·361 | 1,185 | 0·372 |
| Age group 50–64 | 6,400 | 0·199 | 1,038 | 0·210 | 497 | 0·249 | 7,718 | 0·207 | 1,391 | 0·206 | 1,185 | 0·229 |
| Age group 65+ | 6,400 | 0·119 | 1,038 | 0·065 | 497 | 0·062 | 7,718 | 0·144 | 1,391 | 0·064 | 1,185 | 0·058 |
| Higher education | 6,400 | 0·671 | 1,038 | 0·828 | 497 | 0·853 | 7,718 | 0·619 | 1,391 | 0·797 | 1,185 | 0·824 |
| Higher income | 6,063 | 0·425 | 999 | 0·504 | 480 | 0·533 | 6,930 | 0·365 | 1,303 | 0·424 | 1,118 | 0·442 |
| Respondent lives alone | 6,400 | 0·187 | 1,038 | 0·188 | 497 | 0·217 | 7,718 | 0·191 | 1,391 | 0·171 | 1,185 | 0·169 |
| Started to wash hands more often since pandemic | 6,345 | 0·655 | 1,033 | 0·712 | 493 | 0·728 | 7,637 | 0·634 | 1,388 | 0·705 | 1,180 | 0·709 |
| History of illness in the last 3 months | 6,321 | 0·316 | 1,031 | 0·440 | 496 | 0·435 | 7,185 | 0·564 | 1,337 | 0·690 | 1,143 | 0·721 |
| History of COVID-19 testing | 6,400 | 0·152 | 1,038 | 0·225 | 497 | 0·243 | 7,718 | 0·360 | 1,391 | 0·447 | 1,143 | 0·721 |
| Current smoker* | — | — | 949 | 0·205 | 450 | 0·182 | 7,718 | 0·315 | 1,391 | 0·254 | — | — |
| Encouraged to participate in study | 6,400 | 0·231 | 1,038 | 0·232 | 497 | 1·000 | 7,718 | 0·517 | 1,391 | 0·566 | 1,185 | 0·000 |
| CMIA Abbott positive | — | — | 1,038 | 0·093 | 497 | 0·139 | — | — | 1,390 | 0·164 | — | — |
| ELISA Genetico positive | — | — | 1,035 | 0·103 | 495 | 0·147 | — | — | 1,378 | 0·227 | 1,182 | 0·532 |
| ELISA Vector positive | — | — | — | — | — | — | 1,348 | 0·247 | 1,348 | 0·247 | 1,169 | 0·546 |

* — current smoking status variable is gathered from the paper-based survey of tested individuals in the clinic during the first cross-section and is extrapolated for the same individuals for the second cross-section. For the third cross-section all individuals were asked about their smoking status during the phone interview. ** — for the purposes of the analysis we excluded vaccinated individuals from the tested subsample of individuals in the fourth cross-section, assumed that they failed to agree to get tested, and used their predicted seropositivity status from our univariate imputation model rather than the actual test results.

**Table S6.** Representativeness of the survey across study cross-sections

| | Interviewed, 2020-05-25 – 2020-06-28 | Interviewed, 2020-10-12 – 2020-12-06 | KOUZh-2018 |
|---|---|---|---|
| Male, % | 41·2 | 42·5 | 40·2 |
| | (40·0–42·4) | (41·4–43·6) | (38·4–41·9) |
| Age, years | 43·0 | 44·1 | 46·7 |
| | (42·7–43·4) | (43·8–44·5) | (46·1–47·3) |
| 18-34, % | 36·3 | 33·4 | 27·9 |
| | (35·1–37·5) | (32·3–34·5) | (26·3–29·5) |
| 35-49, % | 31·8 | 31·9 | 31·7 |
| | (30·7–32·9) | (30·9–33·0) | (30·0–33·3) |
| 50-64, % | 19·9 | 20·6 | 22·8 |
| | (19·0–20·9) | (19·7–21·5) | (21·3–24·3) |
| 65+, % | 11·9 | 14·1 | 17·6 |
| | (11·1–12·7) | (13·3–14·9) | (16·3–19·0) |
| Education | | | |
| Primary / secondary education, % | 10·0 | 10·5 | 12·9 |
| | (9·2–10·7) | (9·8–11·2) | (11·7–14·1) |
| Special secondary education, % | 23·0 | 27·4 | 39·5 |
| | (22·0–24·1) | (26·4–28·4) | (37·7–41·3) |
| Higher education, % | 67·0 | 62·0 | 47·6 |
| | (65·9–68·2) | (60·9–63·1) | (45·8–49·4) |
| Employed, % | 68·3 | 65·0 | 70·2 |
| | (67·1–69·4) | (64·0–66·1) | (68·6–71·8) |
| Current smoker*, % | 20·5 | 31·5 | 31·6 |
| | (18·0–23·1) | (30·5–32·6) | (30·0–33·3) |
| Self-reported health status | | | |
| Very good, % | 19·9 | 16·2 | 7·8 |
| | (18·9–20·9) | (15·4–17·0) | (6·8–8·7) |
| Good, % | 48·9 | 48·6 | 45·3 |
| | (47·7–50·1) | (47·5–49·7) | (43·5–47·1) |
| Satisfactory, % | 28·5 | 32·3 | 39·2 |
| | (27·4–29·6) | (31·2–33·3) | (37·5–41) |
| Bad, % | 2·4 | 2·4 | 7·1 |
| | (2·0–2·8) | (2·0–2·7) | (6·2–8·0) |
| Very bad, % | 0·3 | 0·5 | 0·6 |
| | (0·2–0·5) | (0·4–0·7) | (0·3–0·9) |
| Lives alone, % | 18·7 | 19·1 | 19·7 |
| | (17·7–19·6) | (18·2–19·9) | (18·3–21·1) |
| Has cellphone, % | 100 | 100 | 99·5 |
| | | | (99·3–99·8) |
| N | 6336 | 7595 | 2977 |

95% confidence intervals in parentheses. “Interviewed” means individuals who agreed to participate in the respective cross-section of the phone survey. KOUZh-2018 is the 2016 round of the Comprehensive Monitoring of Living Conditions household survey carried out by the Federal State Statistics Service of Russia. We subset this survey to include only adults in St. Petersburg. We report only complete-case observations in terms of all variables, therefore the number of observations is slightly lower due to listwise deletion. * — current smoking status variable is gathered from the paper-based survey of tested individuals in the clinic during cross-section 1 (N = 949).

**Figure S1.** Naïve and adjusted seroprevalence by study cross-section and week (ELISA Coronapass)

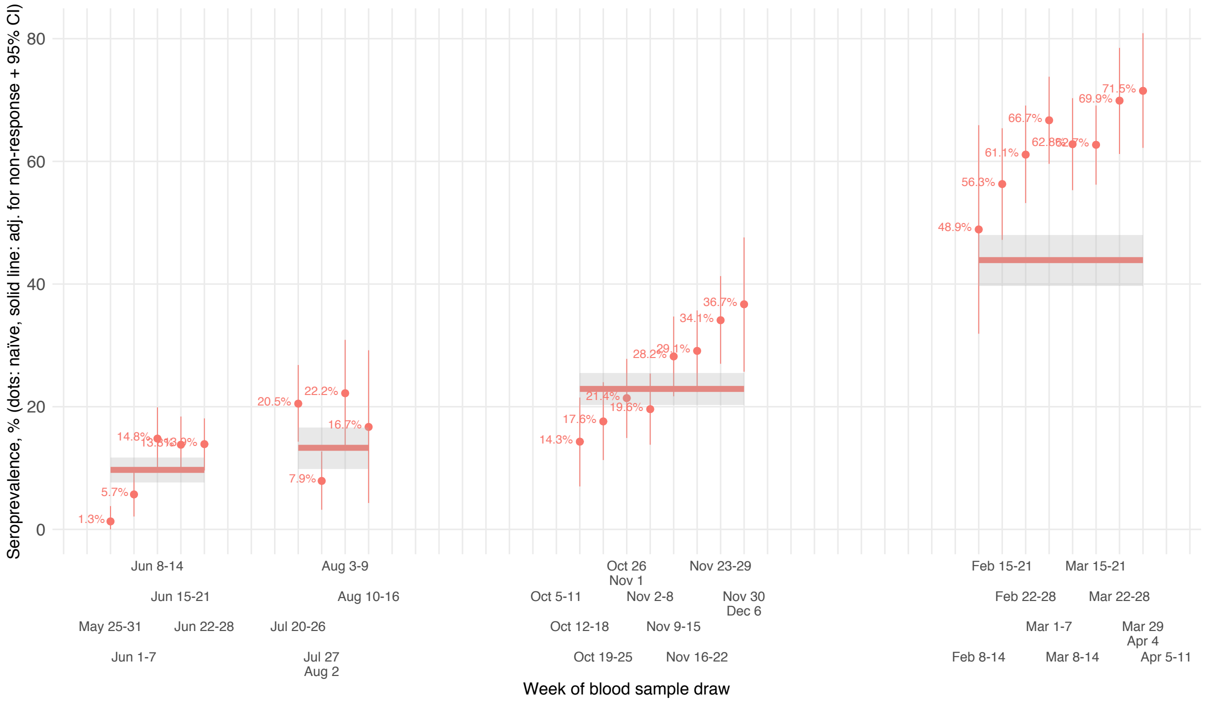

**Table S7.** Seroprevalence by cross-section: naïve adjusted for non-response bias or adjusted for non-response and test performance

| test | CMIA Abbott | | | | ELISA Coronapass | | | | ELISA Vector | | | |
|---|---|---|---|---|---|---|---|---|---|---|---|---|
| estimate | naïve | naïve | adjusted | adjusted | naïve | naïve | adjusted | adjusted | naïve | naïve | adjusted | adjusted |
| raking | no | yes | no | yes | no | yes | no | yes | no | yes | no | yes |
| 2020-10-12 – 2020-12-06 | 14·2 (11·5–16·9) | 14·3 (11·6–17·0) | 11·5 (9·0–14·1) | 11·2 (8·2–14·3) | 11·6 (9·5–13·6) | 11·6 (9·5–13·7) | 9·7 (7·7–11·7) | 9·4 (7·0–11·8) | — | — | — | — |
| 2020-07-20 – 2020-08-08 | 21·1 (16·5–25·7) | 19·7 (15·2–24·3) | 16·1 (11·8–20·4) | 14·0 (9·3–18·6) | 16·5 (13·0–20·0) | 15·5 (12·0–18·9) | 13·3 (9·9–16·6) | 11·4 (7·9–15·0) | — | — | — | — |
| 2020-10-12 – 2020-12-06 | 24·9 (22·0–27·9) | 25·4 (22·4–28·4) | 22·0 (19·0–25·1) | 22·0 (18·5–25·6) | 25·2 (22·8–27·7) | 26·4 (23·9–28·9) | 22·9 (20·3–25·5) | 23·8 (20·7–26·9) | 26·9 (24·3–29·4) | 28·1 (25·5–30·6) | 23·9 (21·3–26·4) | 24·6 (21·5–27·7) |
| 2020-02-15 – 2020-04-04 | — | — | — | — | 57·9 (54·7–61·0) | 56·5 (53·4–59·7) | 43·9 (39·7–48·0) | 42·1 (37·5–46·8) | 58·2 (55·1–61·3) | 57·1 (54·0–60·2) | 49·5 (45·7–53·2) | 48·6 (44·2–52·9) |

### **Table S8.** Seroprevalence by subgroup, ELISA Coronapass

| | | 2020-05-25 – 2020-06-28 | | 2020-07-20 – 2020-08-08 | | 2020-10-12 – 2020-12-06 | | 2020-02-15 – 2020-04-04 | |
|---|---|---|---|---|---|---|---|---|---|
| | | *N* | seroprevalence | *N* | seroprevalence | *N* | seroprevalence | *N* | seroprevalence |
| | | | | | naïve | | | | |
| Age group | 18-34 | 387 | 13·8 (10·2–17·4) | 172 | 16·4 (10·6–22·2) | 499 | 24·4 (20·4–28·4) | 391 | 51·7 (46·3–57·1) |
| | 35-49 | 341 | 9·2 (6·0–12·5) | 158 | 16·5 (10·4–22·6) | 479 | 25·9 (21·7–30·0) | 428 | 61·5 (56·4–66·6) |
| | 50-64 | 199 | 13·7 (8·6–18·7) | 116 | 20·6 (12·9–28·4) | 267 | 28·1 (22·4–33·8) | 257 | 65·1 (58·6–71·6) |
| | 65+ | 61 | 3·6 (0·0–8·4) | 28 | — | 77 | 16·9 (8·1–25·7) | 64 | 42·5 (29·5–55·5) |
| Female | | 623 | 11·0 (8·4–13·6) | 300 | 16·7 (12·2–21·1) | 874 | 23·9 (20·9–26·9) | 747 | 54·0 (50·1–57·9) |
| Male | | 365 | 12·5 (8·9–16·1) | 174 | 16·2 (10·5–22·0) | 448 | 27·9 (23·5–32·3) | 393 | 65·3 (60–70·5) |
| Higher education | no | 168 | 13·6 (8·2–19·0) | 68 | 12·8 (4·5–21·1) | 261 | 28·3 (22·5–34·1) | 200 | 48·9 (41·4–56·4) |
| | yes | 820 | 11·1 (8·9–13·4) | 406 | 17·1 (13·3–21) | 1061 | 24·5 (21·8–27·2) | 940 | 59·8 (56·3–63·2) |
| Higher income | no | 491 | 11·1 (8·2–14·0) | 221 | 13·8 (9·0–18·5) | — | — | — | — |
| | yes | 497 | 12·0 (9·0–15·0) | 253 | 18·9 (13·8–24·0) | — | — | — | — |
| Respondent lives alone | no | 803 | 12·5 (10·1–14·8) | 372 | 18·4 (14·3–22·6) | 1094 | 26 (23·3–28·8) | 951 | 59·8 (56·3–63·2) |
| | yes | 185 | 7·6 (3·6–11·6) | 102 | 9·6 (3·6–15·6) | 228 | 21·5 (15·8–27·1) | 189 | 48·3 (40·6–56·0) |
| History of COVID-19 testing | no | 760 | 8·4 (6·4–10·5) | 357 | 11·6 (8·1–15·0) | 726 | 16·2 (13·4–19·0) | 319 | 35·4 (29·8–41·0) |
| | yes | 228 | 21·9 (16·3–27·6) | 117 | 31·6 (22·6–40·5) | 596 | 36·3 (32·2–40·4) | 821 | 66·6 (63–70·2) |
| Current smoker | no | — | — | — | — | 982 | 27·2 (24·3–30·2) | — | — |
| | yes | — | — | — | — | 340 | 19·5 (15·1–23·9) | — | — |
| History of illnesses in the past 3 months | no | 547 | 5·2 (3·2–7·1) | 266 | 9·8 (6·1–13·5) | 407 | 11·8 (8·5–15·0) | 319 | 35·4 (29·8–41·0) |
| | yes | 441 | 19·5 (15·6–23·4) | 208 | 25·1 (18·9–31·3) | 915 | 31·2 (28·1–34·4) | 821 | 66·6 (63·0–70·2) |
| Started to wash hands more often since pandemic | no | 279 | 16·4 (11·8–20·9) | 128 | 24·6 (16·7–32·5) | 389 | 26·0 (21·4–30·6) | — | — |
| | yes | 709 | 9·7 (7·4–11·9) | 346 | 13·5 (9·7–17·3) | 933 | 24·9 (22·0–27·9) | — | — |
| | | | | | adjusted for non-response bias and test characteristics | | | | |
| Age group | 18-34 | 387 | 12·1 (8·7–15·5) | 172 | 14·0 (8·8–19·2) | 499 | 23·2 (19·4–27·0) | 391 | 41·5 (36·1–47·0) |
| | 35-49 | 341 | 8·0 (5·1–10·8) | 158 | 15·5 (9·7–21·3) | 479 | 24·1 (20·2–28·1) | 428 | 48·1 (42·4–53·8) |
| | 50-64 | 199 | 11·6 (7·1–16·0) | 116 | 15·5 (8·7–22·3) | 267 | 25·0 (19·8–30·2) | 257 | 50·4 (43·1–57·6) |
| | 65+ | 61 | 3·3 (0·0–7·7) | 28·0 | — | 77 | 15·9 (7·8–24) | 64 | 29·5 (18·4–40·5) |
| Female | | 623 | 9·3 (7·0–11·7) | 300 | 14·2 (10·2–18·2) | 874 | 22·1 (19·1–25·1) | 747 | 37·9 (33·4–42·4) |
| Male | | 365 | 10·2 (7·1–13·4) | 174 | 11·9 (7·1–16·8) | 448 | 23·9 (19·9–27·9) | 393 | 52·0 (46·0–57·9) |
| Higher education | no | 168 | 10·5 (6·2–14·7) | 68 | 8·8 (2·7–14·8) | 261 | 24·1 (19·0–29·2) | 200 | 37·3 (30·2–44·4) |
| | yes | 820 | 9·4 (7·3–11·4) | 406 | 15·4 (11·6–19·2) | 1061 | 22·1 (19·6–24·7) | 940 | 47·5 (43·2–51·8) |
| Higher income | no | 491 | 9·2 (6·6–11·8) | 221 | 11·1 (7·1–15·2) | — | — | — | — |
| | yes | 497 | 10·4 (7·6–13·2) | 253 | 16·1 (11·2–20·9) | — | — | — | — |
| Respondent lives alone | no | 803 | 10·6 (8·3–12·8) | 372 | 14·6 (10·7–18·4) | 1094 | 23·8 (21·0–26·6) | 951 | 45·5 (41·0–49·9) |
| | yes | 185 | 5·9 (2·6–9·2) | 102 | 7·4 (2·6–12·2) | 228 | 18·7 (13·8–23·7) | 189 | 36·9 (29·8–44·0) |
| History of COVID-19 testing | no | 760 | 7·7 (5·6–9·7) | 357 | 10·2 (6·8–13·5) | 726 | 15·2 (12·3–18·1) | 319 | 34·3 (28·8–39·9) |
| | yes | 228 | 20·9 (15·4–26·3) | 117 | 30·2 (21·3–39·1) | 596 | 36 (31·8–40·3) | 821 | 66·5 (62·7–70·3) |
| Current smoker | no | — | — | — | — | 982 | 24·8 (21·9–27·8) | — | — |
| | yes | — | — | — | — | 340 | 18·5 (14·3–22·8) | — | — |
| History of illnesses in the past 3 months | no | 547 | 5·4 (3·3–7·5) | 266 | 9·0 (5·4–12·7) | 407 | 11·7 (8·3–15·2) | 319 | 39·6 (34·8–44·5) |
| | yes | 441 | 18·9 (14·9–22·8) | 208 | 22·3 (16·3–28·2) | 915 | 31·5 (28·1–34·9) | 821 | 49·1 (45·7–52·6) |
| Started to wash hands more often since pandemic | no | 279 | 12·7 (8·8–16·5) | 128 | 19·2 (12·6–25·8) | 389 | 24·0 (19·6–28·4) | — | — |
| | yes | 709 | 8·2 (6·1–10·3) | 346 | 10·2 (6·9–13·5) | 933 | 22·2 (19·3–25·1) | — | — |
